# Ensemble Forecasts of Seasonal Dengue Epidemics

**DOI:** 10.1101/2021.03.09.21253185

**Authors:** Yuliang Chen, Tao Liu, Sen Pei, Xiaolin Yu, Qinghui Zeng, Haisheng Wu, Jianpeng Xiao, Wenjun Ma, Pi Guo

## Abstract

As a common vector-borne disease, dengue fever still remains a lot of challenges to forecast for which the significant distinction of epidemic scale is affected by multiple factors, such as mosquito density, meteorological conditions, geographical environment, travel and so on. To track down the epidemic scale and forecast the remaining time of epidemic season, the population size affected by the epidemic is evaluated before the compartmental model is optimized by assimilation observation with filtering method. In retrospective forecast of dengue pandemic for Guangzhou from 2014-2015 seasons, accurate forecast of dengue cases is generated with an accurate prediction of peak time in all time periods. The real-time forecast system shows a good performance on capturing the trajectory of dengue transmission and scale of epidemic.

## Introduction

Dengue fever, as one of the key neglected tropical diseases (NTDs), got attention from World Health Organization (WHO) ^1^, threatens the health of billions of people in the world. More than 390 million people are infected with dengue every year ^2, 3^. About 75% of dengue burden is concentrated in Southeast Asia and the Western Pacific near the tropics. Guangdong province is one of the most serious prevalent areas in China. During the period of 2011-2017, dengue fever cases in Guangdong occupied 81% of the total in mainland China, especially in Guangzhou city, accounting for 76% of Guangdong (Figure 1a). In addition, lengthened with the warm season owing to global warming in recent years, mosquitoes have become more active that potentially increasing the risk of Mosquito-borne diseases outbreaks ^4, 5^.

**Figure 1.**
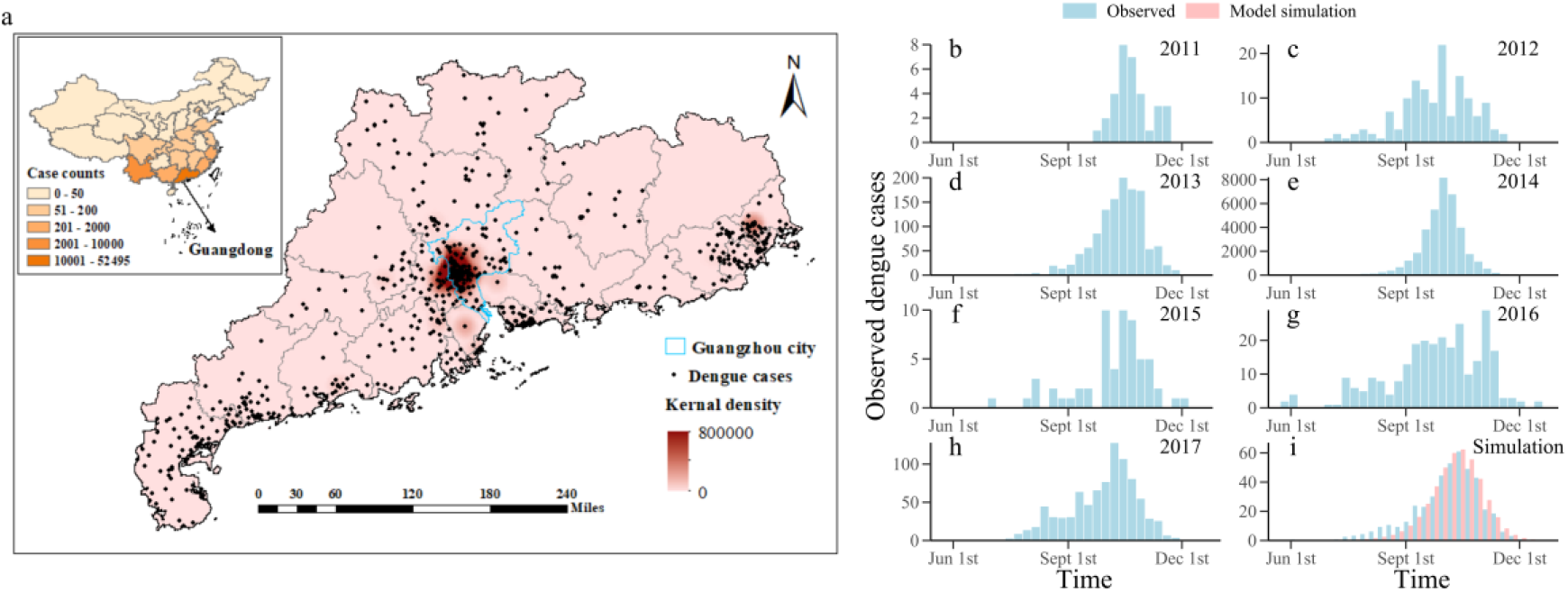
Seven consecutive dengue seasons (2011-2017) weekly observations and its geographical distribution, and free model simulation of dengue cases. **a** Geographical distribution of dengue cases and its kernel density estimation in Guangdong Province over the study period from January 2011 to December 2017. Cases counts and distinguished by colored according to the magnitude in each province also present (inset of **a**). In these 7 years, the cumulative number of dengue cases in Guangdong province accounted for 81% of the China mainland, of which Guangzhou took account for 76% of Guangdong province. **b-h** Weekly observations (from Jun 2011 to Dec 2017) of dengue cases in Guangzhou. **i** Average weekly observations (2011-2017, blue bar) and free model simulation (pink bar) of dengue cases. Only 35% of human cases had been reported around the season.

There was a series of outbreaks of dengue fever in China during 2014, with the number of infected people reaching 46,864 ^6^, which substantially threatened the public health. However, affected by community prevention and different government’s mosquito control strategy each year, the measures in this regard will bring uncertainty to the prediction. Furthermore, the main characteristics of dengue fever prevalence and transmission in China are as follows: (1) the intensity of dengue fever outbreak varies from year to year, for example, the weekly number of new infected dengue cases in Guangzhou city is sometimes less than 10 cases and sometimes more than 8,000 cases at peak; (2) there are generally no cases or only one or two cases of dengue fever in other time except for outbreak (as shown in Figure 1b-h, the number of dengue fever cases in non-outbreak period was basically zero around the year). We can observe that the time series of dengue onset contain a large number of zero values per unit time, and the data with zero-inflated characteristics exceed the predictive power of general discrete distributions such as Poisson distributions ^7^. If the peak intensity and peak time of dengue fever can be accurately predicted, it will be conducive to the rapid response of government and strengthen the elimination of mosquito breeding places in the outbreak areas.

However, there is still immature to forecast on dengue pandemic, especially for China area. Basically, current forecast methods of infectious diseases concentrate on three kinds of categories: statistical approaches (e.g., time series analysis, Bayesian modeling averaging ^8-11^), state space estimation methods (e.g., model-inference systems ^12-15^) and deep learning (neural network algorithm^16^). Recently, a growing number of forecast researches developed different model-inference frameworks and used them to generate accurate ensemble forecasts. Coupling with data assimilation algorithm, the dynamic model can be coupled with the observed data stream and bring out the real-time prediction of infectious diseases, such as West Nile virus (WNV), Ebola and influenza ^17-21^.

This study will extend the algorithms mentioned above to generate a real-time forecast of dengue fever. Initially, the dengue dynamic model of mosquito-borne transmission was constructed, and then coupled with the observed stream. In particular, reported dengue cases were assimilated using the ensemble adjusted Kalman filter (EAKF) ^22^. Given the consequences caused by different intensity of outbreak, we suggest using a self estimating population size approach embedding in each data assimilation step. Retrospective forecasts of dengue were performed and an accurate real-time prediction was made for the 2011-2018 dengue seasons using the above prediction framework.

## Results

### Distribution of dengue cases

During 2011-2017, Guangdong province accounted for 81% of China’s total reported dengue cases (Taiwan province was not included in the report), among which Guangzhou city was the most serious epidemic area, accounting for 76% of the total reported dengue fever cases in Guangdong Province. According to the spatial distribution of dengue cases in 2014 shown in Figure 1a, dengue cases were mainly distributed in East and West of Guangdong, Pearl River Delta and other densely populated areas with developed water system. In particular, the Pearl River Delta was the most serious epidemic area of dengue fever in that year, and Guangzhou (blue area) was located in the center of the disaster area. In this study, a real-time operational forecast was generated with timely reported dengue cases of Guangzhou from 2011 to 2017.

However, it was different between each scale of annual dengue outbreak. As shown in Figure 1b-h, although dengue fever epidemic in seven years had similar seasonal outbreak characteristics, the scale of dengue outbreak was obviously different, with 8000 cases in 2014 while less than 10 cases in 2011. Because the general SIR-EAKF system ran by using a fixed population size, it was unable to catch the structure of pandemic when the population size was set lower than the true value for the excessive consumption of the susceptible, resulting in a sharp decline in the subsequent prediction accuracy. Simultaneously, if the population size was set much larger than the true value, the SIR-EAKF system would depend on the simulation with a high value owing to large population size rather than the observation. It would take a long time for assimilation to capture the trajectory of the low outbreak year, so it was unable to get an accurate forecast.

### Construction of SIR-EAKF and simulation of synthetic outbreak

In order to better reproduce the actual transmission path of dengue fever, the dengue SIR model constructed in this study could simulate the horizontal transmission of dengue virus between human and mosquito ^23^ and the vertical transmission of mosquito. Using the information of mosquito density and ambient temperature, the annual average time series of mosquito natural birth rate and mosquito transmission probability were calculated, respectively and were imported in the compartmental model, to simulate the seasonal epidemic characteristics of dengue fever. Figure 1i shows the free simulation of the annual dengue case from 2011 to 2017 (except 2014) by the dengue SIR model. It can be seen that the dengue SIR model fits the historical data well.

We used the dengue SIR model to get the simulated dengue case through free simulation with historical data. The simulated dengue case was taken as the true value and was used to test the adjustment of EAKF on state variables and model parameters. Throughout our study, all real-time forecasts were made by a system that integrating SIR-EAKF with an automatic method for determining the size of epidemic-affected population, which was referred to the combined SIR-EAKF model. Supplementary Figure 1 shows the time series results of posterior ensemble for state variables and model parameters generated using the combined SIR-EAKF model. For state variables *I*_*M*_, *I*_*H*_, *NewI*_*M*_ and *NewI*_*H*_, the model can stably capture the trajectory in the whole prediction process. Affected by the self-determination method, the prediction of state variables *S*_*M*_ and *S*_*H*_ are lower than the true. It is inaccurate for *N* the same as *S*_*M*_ and *S*_*H*_ until assimilation more observations to the 20th week. However, the estimation of model parameters, *D* and *β*, are basically adjusted to the true and fluctuate around it.

### Stability of the combined SIR-EAKF system

In order to test the stability of the combined SIR-EAKF system, sensitivity analyses were performed on time interval, ensemble number and OEV ^19^ (Figures 5-7). Time interval sensitivity analyses of synthetic truth were run by the combined SIR-EAKF using 300 ensemble members.

Supplementary Figure 5 shows that when the time interval is shorter, the model can better capture the real value of sum. When the time interval increases, the interval of prediction set increases correspondingly, and the uncertainty of prediction increases. Supplementary Figure 5 shows that the EAKF can better align to the true of *I*_*M*_ and *I*_*H*_ with time interval decreasing. When the time interval increases, the inter quartile range (IQR) of ensemble simulation increases correspondingly with the uncertainty of prediction. Adjustment of model parameters *D* and *β* are both performed well in different time interval situations. But for state variables *S*_*M*_ and *S*_*H*_, the longer the time interval is, the smaller the IQR of ensemble simulation is. It may be due to the decrease of observation assimilation times and the corresponding decrease of variance expansion times. The overall time interval sensitivity analysis shows that the combined SIR-EAKF framework has stable prediction and correction ability under different time interval settings.

In the same way, the synthetic true is used to conduct sensitivity analysis on the set of ensemble number. The combined SIR-EAKF model was run for 250 times to observe the distribution of mean ensemble estimator for state variables and model parameters in the 22nd week, as shown in Supplementary Fig. 6. The certainty of prediction enhances with the increase of ensemble number, which also resulting in the increase of computation. Consequently, considering the balance between the certainty of prediction and computation, the number of ensemble was set as 300 in this retrospective analysis. In the OEV sensitivity analysis, the results are shown in Fig. 7. We used different values to multiply OEV to verify the dependence of the combined SIR-EAKF model on OEV. The results show that with the increase of OEV, the EAKF assimilation tends to follow the model simulation and deviates from the observation when the variance of ensemble is smaller than OEV. However, with the decrease of OEV, the variance of ensemble simulation is larger than the OEV during initial assimilation period. Thus, a greater weight on observation is given to make the ensemble simulations shrinking near the observation during the assimilation by EAKF, reducing the uncertainty of forecasts.

### Retrospective forecasts

Next, we used the combined SIR-EAKF system to generate a retrospective weekly forecast of dengue cases in Guangzhou from 2011 to 2017. In order to reduce the effect of filtering divergence, the forecast system starts from the 20th week of each year and ends at the 19th week of next year. By integrating observation stream data and simulation data, the EAKF can better estimate ensemble state variables and model parameters until the end of the epidemic season. Figure 2a shows the successive forecasts of dengue cases during 2013-2014 seasons by the combined SIR-EAKF. As can be seen from the figure, the trajectory of outbreak was better aligned before reaching the peak (the 23rd week of the epidemic season). With more observation assimilation, the ensemble range was further reduced. In addition, Figure 2b also shows the distribution of ensemble prediction of peak time. The predicted average peak time of dengue fever was within ± 1 of the actual peak week. Because dengue fever is an epidemic with strong seasonality, our SIR model also has strong seasonality, which can accurately predict the peak of dengue fever outbreak. In other words, the seasonal feature of dengue epidemic is accurately reflected by our dengue SIR model.

**Figure 2.**
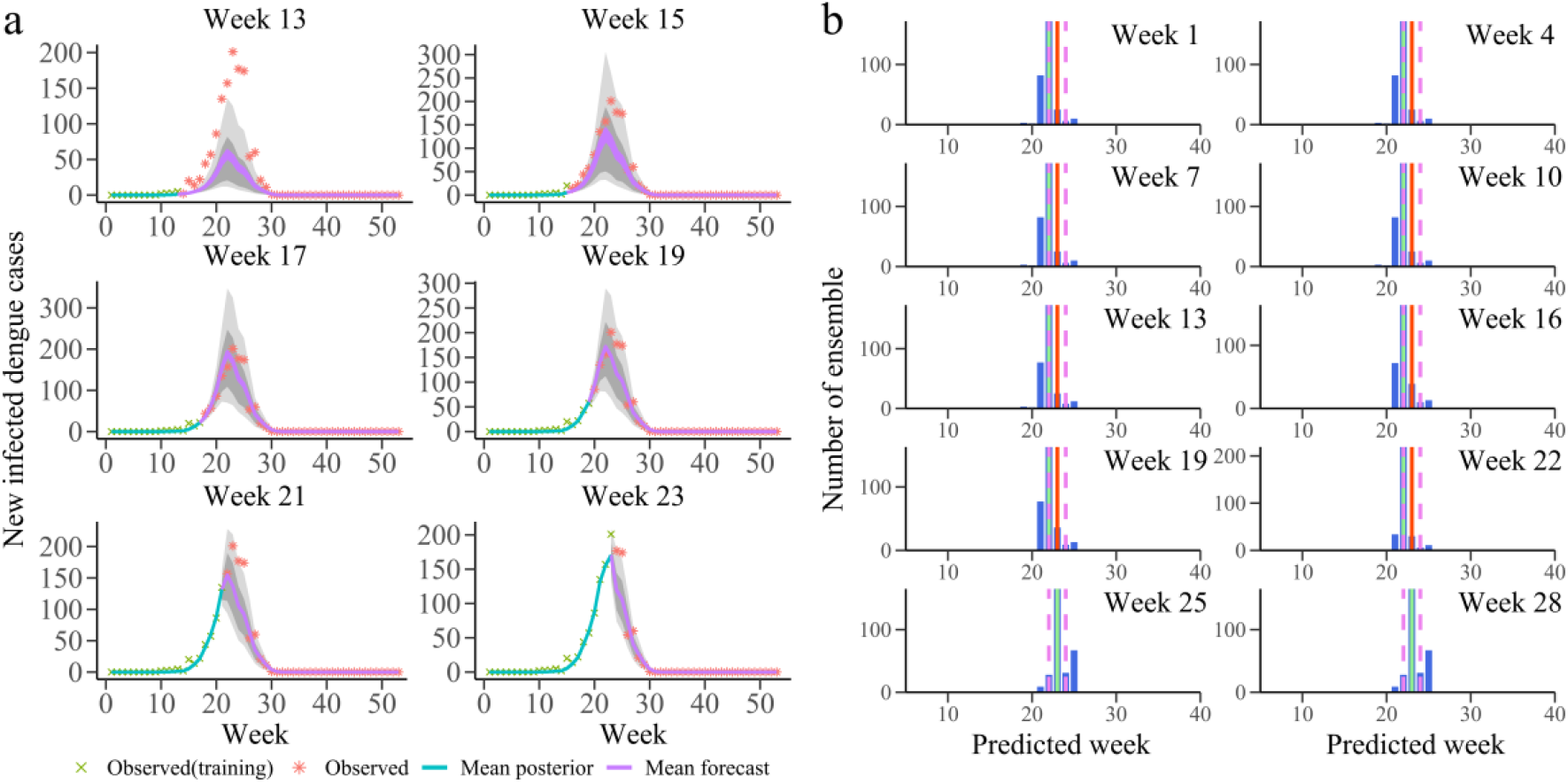
Results of 300-member SIR-EAKF retrospective forecasting for 2013-2014 season. **a** Ten bi-weekly forecast of dengue cases for2013-2014 season. The purple lines are the ensemble mean forecasts, the grey area is the spread of the ensemble forecast (light grey represents area between 10th and 90th percentile and the darker grey area represents the spread between the 25th and 75th percentile), blue lines are the ensemble mean posterior distribution, green x are data points assimilated into the model and red * are future observations. **b** Histogram of ensemble forecast peak timing for prediction initiated at the end of weeks 1, 4, 7, 10, 13, 16, 19, 22, 25 and 28 week (blue). Also shown are the observed peak (red, week 23) and its ±1 week (pink) and the ensemble mean (green).

Afterwards we evaluated retrospective forecast for seven seasons using three metrics ^17^: peak time, peak intensity and total incidence. Each metric is deemed accurate if: the predicted peak time is within ± 1 week of the observed peak time; the predicted peak intensity is within ± 25% (± 1 case) of the observed peak intensity; the predicted total incidence is within ± 25% (± 1 case) of the observed total incidence. Based on these conditions, the evaluation of three indicators for seven dengue seasons is shown in Supplementary Figures 2-4. It can be found that the prediction of peak time has a good performance in the whole prediction process. Besides, the prediction of the peak intensity and total incidence can give an accurate prediction one week ahead of the peak time.

Figure 4a-c shows the ensemble distribution of the distance between prediction and observation of the three metrics for seven dengue seasons. As a result, most ensembles can accurately predict the peak time ahead of the observed peak time. For the peak intensity and the total incidence, with the accumulation of assimilation, ensemble distribution of the log(difference) gradually approaches zero when the predicted time is at or past the observed peak time. Finally, the overall prediction accuracy evaluation fraction of the combined SIR-EAKF model for three metrics in seven epidemic seasons is given (Figure 4d). The forecast is grouped according to the predicted lead time, which is defined as the week of forecast generation minus the week of observed peak weekly dengue reported cases. Overall, forecasts of peak time were accurate > 87.5% of the time with 1 week prediction lead. Forecasts of peak intensity were also > 75% with one with ahead the observed peak time. For the total incidence, there are 62.5% of forecasts were accurate 1 week before the observed peak time, and > 75% of forecasts were accurate 2 week after the observed peak time.

**Figure 4.**
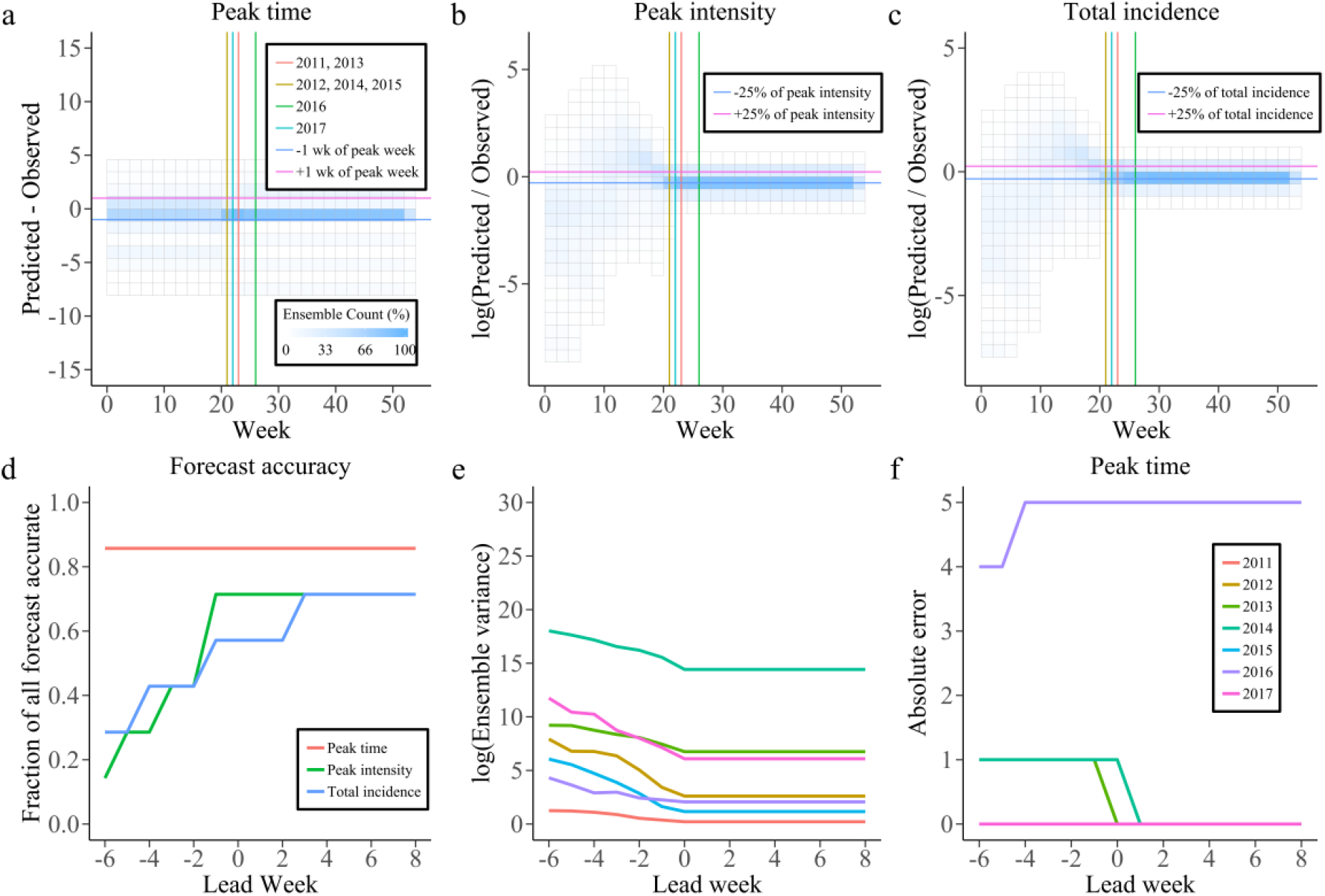
Results for 2011-2017 retrospective forecasts. **a-c** Distributions of the peak timing (peak intensity and total incidence) distance from the predicted values to the observed target on each predicted week. The peak time for different dengue season is distinguished with distinct color vertical lines. The horizontal lines are representing the +1 wk (+25%, pink) and the-1wk (−25%, blue) from the observed peak week (peak intensity or total human dengue cases).**d** The fraction of forecasts accurate as a function of lead week for the metrics peak timing (red), peak intensity (green) and total incidence (blue). A forecast can be considered as accurate when (1) peak timing was within ± 1 week of the observed peak of weekly new infected dengue cases; (2) peak intensity was within ± 25% or ± 1 cases of the observed peak weekly new infected dengue cases; (3) total incidence was within ± 25% of the observed. **e, f** characteristics the ensemble variance (**e**)and absolute error (**f**) for those predictions of each pandemic seasons which distinguished with different color.

In addition, Figure 3a illustrates the effectiveness of the self-determination method. With the increase of assimilation observational data, the population size affected by the epidemic of each dengue season can be well predicted before the peak time. It is slightly higher than the cumulative number of dengue cases for estimation, providing a sufficient and appropriate fixed population size for the general SIR-EAKF. As shown in Figure 3c, except for the low outbreak season, the posterior ensemble mean estimated by the combined SIR-EAKF model can well predict the dengue outbreak. Moreover, the analyses of ensemble variance grouped by lead time (Figure 4e) depict that there is a downward trend for ensemble variance with more observations assimilation. Figure 4f shows the absolute difference between the ensemble average peak time and the observed peak time in seven epidemic seasons grouped by lead time. Except for the abnormal peak time in the 2016-2017 dengue seasons, other dengue seasons can accurately forecast the peak time 6 or more consecutive weeks in advance.

**Figure 3.**
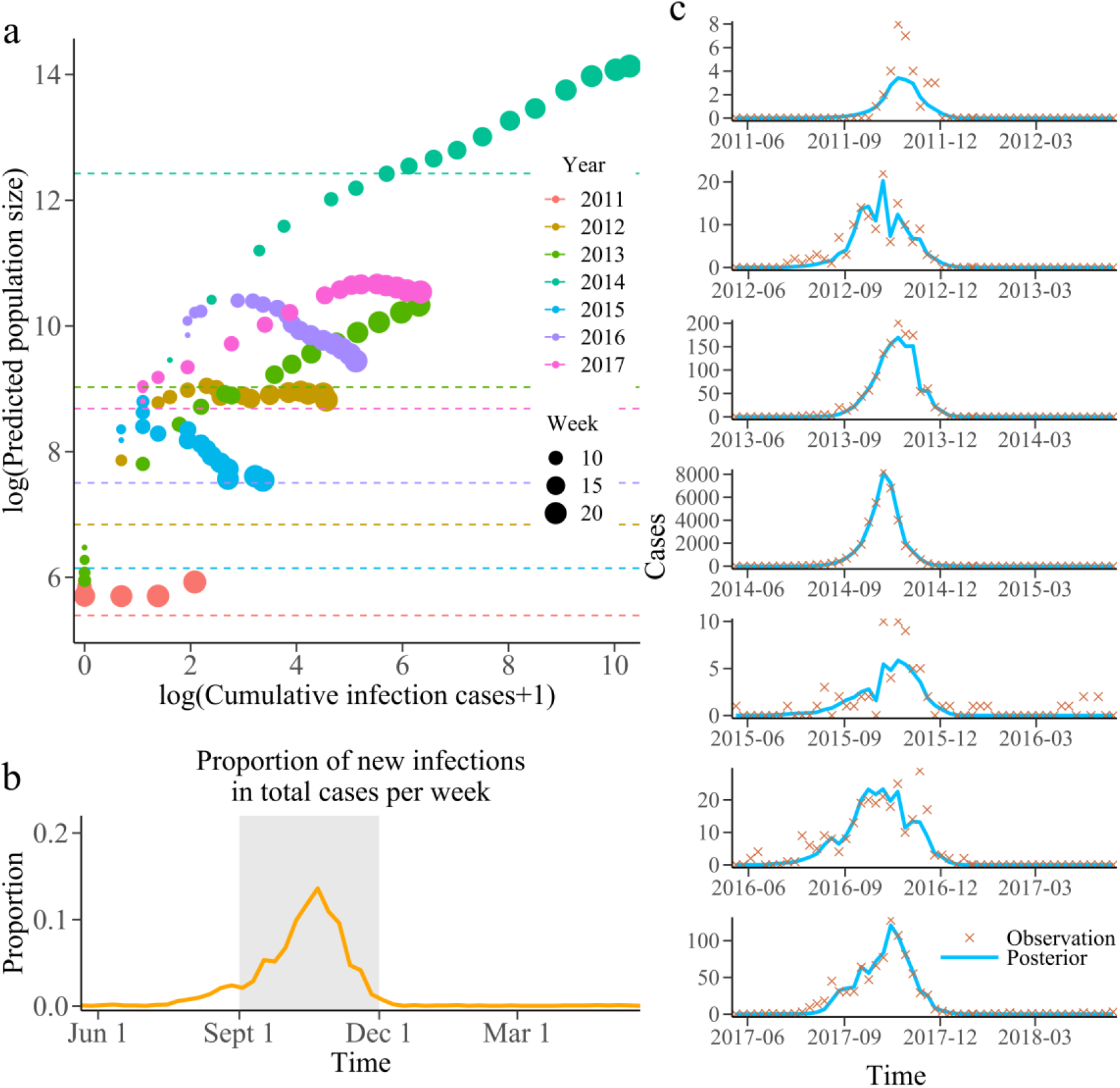
Prediction of population size and its function. **a** The population size set within the 5thand 22nd (the average peak time) weeks of each year. Each color represents corresponding year and the circle size indicate the predicted week. The dotted lines represents log(cumulative dengue cases over the corresponding season). The abscissa is log(cumulative infection number from the beginning of the dengue season to the current week +1). Besides, the ordinate is log(predicted population size).Based on the cumulative infection number from the beginning of the season to the current week, the total infection number was estimated as the population size of SIR-EAKF. For the convenience of display, log exchange was performed here. **b** Average proportion of new infections in total cases per week. The grey area shown that these 13 weeks (from 15 to 28 week) contain 90.7% of the total cases in the whole season except 2014-2015 season. **c** Weekly observed human dengue cases (cross symbols) for each year. The solid blue lines are the posterior mean of the SIR-EAKF fit.

## Methods

### Construction of dynamic model of dengue fever transmission

In our study, to simulate dengue outbreak dynamics, a standard susceptible-infected-recovered (SIR) epidemiological model modulated by local ambient temperature conditions is developed. Besides, the transmission of dengue fever between mosquitoes and humans is also described in the mathematical model ^24^. Assuming a perfectly mixed population, the compartmental model describing the dynamics of dengue transmission can be constructed by the following equations.

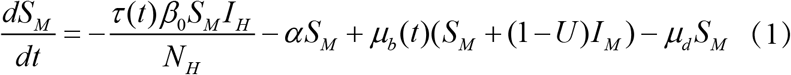

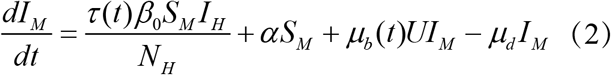

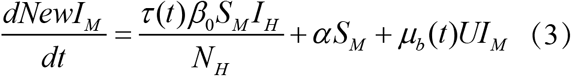

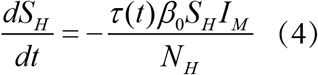

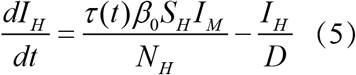

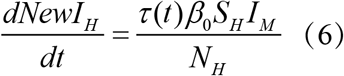

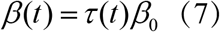

Where *S*_*M*_ is the number of susceptible mosquitoes, *I*_*M*_ is the number of infected mosquitoes, *NewI*_*M*_ is the number of new infected mosquitoes, *S*_*H*_ is the number of susceptible humans, *I*_*H*_ is the number of infected humans, *NewI*_*H*_ is the number of new infected human, *N*_*M*_ is the mosquito population, *N*_*H*_ is the human population, *α* is the rate of dengue seeding into the local model domain, *U* is the dissemination rate and is constant over an outbreak, *μ*_*b*_ (*t*) is the mosquito birth rate at time *t, μ*_*d*_ is the mosquito death rate and is constant over an outbreak, *τ* (*t*) is the population transmission rate at time *t, β*_0_ is the basic contact rate between humans and mosquitoes, *β* (*t*) is the contact rate between humans and mosquitoes at time *t, D* is the mean infectious period for human.

Here, to construct the *μ*_*b*_ (*t*) of compartmental model, MOI information is converted into mosquito natural birth rate. And the population transmission rate at time *t, τ* (*t*), is translated from the ambient temperature using linear regression piecewise function according to the previous study ^25^. It can be seen from Supplementary Information for more details on the calculation of *μ*_*b*_ (*t*) and *τ* (*t*). Throughout the study, we set a constant population of human with no birth or death and time-variant population of mosquitoes with the seasonal natural birth ^26^. Besides, we consider vertical transmission of mosquitoes using a constant dissemination rate, *U*, to develop the transmission path of dengue fever. In addition, the simulation was seeded with infected mosquitoes at a rate of 1 in 500,000 susceptible mosquitoes^17^. For model scaling, we assumed that the number of dengue cases reported in clinics represent 35% of total new infections each week. Following Eqs. (1)-(7) mentioned above, the compartmental model is then integrated forward using classical Runge-Kutta method, which can provide a more accurate prediction for state variables in each forecast.

### Observational data

The individual surveillance data of dengue fever cases in Guangzhou from 2011 to 2018 were obtained from Guangdong Provincial Centers for Disease Control and Prevention (CDC). All human dengue cases were diagnosed according to the diagnostic criteria for dengue fever (WS216-2008) enacted by Chinese Ministry of Health ^27, 28^. Among them, dengue virus subtypes were divided into four types, all of which were included in the statistics. Only the local cases were included in this study to avoid the uncertainty of import cases. Dengue cases were aggregated by week according to the date of illness onset with each week defined as Sunday to Saturday. Figure 1b-h shows the weekly observations of dengue cases from Jun 2011 to Dec 2017 in Guangzhou.

Mosquito Oviposition Index (MOI), a kind of surveillance data of mosquito density, in Guangzhou from 2011 to 2018 were obtained from Guangdong CDC. The MOI is defined as number of ovitrap with positive adult and egg of Aedes albopictus / number of effective ovitrap ^29^. The annual series of MOI of 2011 to 2018 is used to indicate the natural growth of mosquito population size. Assuming that the natural growth of mosquito population size in each year is zero, that is to say the population size of mosquito is equal at the same time in each year. The annual series of MOI can convert into the natural birth rate of mosquito, which is then used to calculate the *μ*_*b*_ (*t*) and *μ*_*d*_ of dengue compartment model (see the Supplementary Information for more details).

Daily meteorological data on ambient temperature, maximum temperature and rain volume for the same period were publicly available on the China Meteorological Data Sharing System (http://data.cma.cn/). Several studies have shown the association between the reproduction of dengue virus in mosquitoes and the ambient temperature ^25,30^. Based on previous studies, a function was constructed to represent the relationship between ambient temperature and population transmission rate, namely the transmission probability of dengue virus by mosquito bites *τ* (*t*) (see Supplementary for more detail on the construction of this function).

### Compartmental model-EAKF framework

Previous studies ^17, 19, 31^ have used the EAKF assimilation method in conjunction with a variety of compartmental epidemiological models to assimilate the observation data and update the simulation data based on Bayes" rule. EAKF adjusts ensemble of model simulation state variables to true state. Using cross ensemble co-variability, unobserved state variables and parameters are also updated. For further details of EAKF algorithm, see Anderson ^22^. In this study, a 300-member ensemble simulation of the SIR compartmental model was run in conjunction with the Guangzhou dengue cases data and the EAKF. There were 6 state variables and 3 parameters *X*_*t*_ = (*S*_*M*_, *I*_*M*_, *S*_*H*_, *I*_*H*_, *D, β, N, NewI*_*M*_, *NewI*_*H*_) and weekly observations of human dengue cases 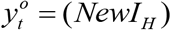 included in the filtering frame work to estimate the true state and parameter of system.

The model assumed that the mosquito population size was time-varying for the combined affection of the seasonal birth rate and constant death rate. Besides, the scaling of observed human dengue cases to the model simulation was assumed to be one. The EAKF consists of 300-member ensemble of SIR model replicates each of which is initialized from a randomly drawn suite of state variable conditions and parameter values. The ensemble dengue SIR model was used to predict the next state variables and then updated with EAKF on observation state variables. In addition, the inter-variable relationships were assumed to be linear. Consequently, a cross ensemble co-variability method was used to adjust the unobserved state variables and parameters (*S*_*M*_, *I*_*M*_, *S*_*H*_, *I*_*H*_, *D, β, N, NewI*_*M*_) by multiplying the ensemble covariance with the observation adjustments in EAKF algorithm. The SIR-EAKF model was then integrated forward to the next observation using the update (posterior) state variables and parameters and the data assimilation updating process is repeated. Over time, the observed dengue cases were used for recursive adjustment to optimize the model state variables and parameters, so that the integrated model simulation can better simulate the local outbreak dynamics.

If the ensemble spread tend to too small due to long time iteration of filter adjustment, it will cause the EAKF system diverge from the true trajectory, which is called "filter divergence". In order to avoid filter divergence, multiplicative inflation factor *λ* =1.025 was multiply with the prior ensemble before assimilation. Besides, inspired by previous studies, we use a heuristic observational error variance (OEV) in running the EAKF, which consists of a baseline uncertainty and a proportional part determined by the new infection dengue cases on that week. Specifically, the OEV for week *t* was *OEV*_*t*_ = (*Obs*_*t*_^2^ +100) / 25.

### Size (*N*) of epidemic-affected population and the combined SIR-EAKF

Due to the huge difference between peak intensity of each year, it is irrational to give a too large fixed *N* for a dengue season with small peak intensity, and the same applied to give a too small fixed *N* for a dengue season with large peak intensity. Further discussion of this problem can see the Supplemental Information for more detail. Consequently, we consider the population size affected by the epidemic that varies with the observed time series of dengue cases to solve this problem.

In this study, in order to solute the problem of different scales of outbreaks in each epidemic season, we suggest to use a simple self-determination method for the *N*. The population size affected by the epidemic, *N*, was calculated from the historical data and used as a fixed parameter of SIR-EAKF. It should rerun the SIR-EAKF model to assimilate observed data when the affected population size was updated. In particular, first of all, the average proportion of weekly new infected dengue cases in the total number of infected dengue cases in that year was calculated according to the history data from 2011 to 2017 (except year of 2014 which have an abnormally high value; it can be seen from Figure 4b.). After that, the value of *N* in current time was estimated through the average proportion sequence and the current observation. By estimating and updating *N* (the fixed parameter of SIR-EAKF) at time *t*, the assimilation and update process was needed to rerun the time period of {1, 2,…,*t*}. Moreover, in the combined model, this method was set to run from the fifth week to the average peak time of 22 weeks to reduce the amount of computation, and the lower limit of the population size affected was 300 and the upper limit was 1,500,000.

### Retrospective forecasts

Guangzhou is one of the most prevalent areas of dengue fever in Guangdong Province, accounting for 78% of cumulative dengue cases reported from 2011 to 2018 in Guangdong Province. Therefore, Guangzhou is a good representative for analyzing the development of dengue fever of Guangdong Province. We used an ensemble compartmental-model initiated with a 300-member ensemble to retrospectively forecast Guangzhou"s dengue epidemic for outbreak season of 2011 to 2017. Since the outbreak of dengue starts in June and ends in December of that year, in order to reduce the filter divergence caused by long time adjustment of EAKF, a dengue season is moved forward and is defined as the 20th of week of each year to the 19th week of next year in running SIR-EAKF system. Each ensemble member state variables was initialized with: *S*_*M*_ (0) = *N*_*M*_ (0) − *I*_*M*_ (0), *I*_*M*_ (0) = *U*(0, *N*_*M*_ /1000), *S*_*H*_ (0) = *N*_*H*_ ×*U* (0.7, 0.9) − *I*_*H*_ (0), *I*_*H*_ (0) = *U* (0,1) ; and model parameters were randomly selected from uniform distributions: *D* = *U* (5, 7), *β*_0_ = *U* (0.045, 0.055), *N* = *N*_*H*_ = *N*_*M*_ (0) = *N*_*p*_ / *U* (0.6, 0.8), and average of peak time was set as the 22th week of the dengue season. The simulation was seeded with infected mosquitoes, *α*, during integration over season at a rate of 1 in 500,000. In addition, each ensemble member with the constant model parameter: *U* = 0.25 and *μ*_*d*_ = 1/ 15 over an outbreak.

After retrospective forecast for each dengue season, we analyzed the accuracy of ensemble forecasts and compared with observations using peak timing, peak magnitude and total magnitude of the seasonal number of dengue cases ^14, 17^. For all three metrics, we compared the ensemble mean trajectory with observation. An accuracy ensemble forecast should meet the following conditions: 1) it peaked within ±1 week of the observed peak of new infected dengue cases; 2) the maximum new infected dengue cases was within ±25% or ±1 of the observed peak intensity; 3) the total number of new infected dengue cases over the entire season was within ±25% or ±1 of the observed, whichever was larger. Besides, all forecasts were grouped by the same prediction lead and the fraction of accurate forecasts was quantified. In particular, the prediction lead means that how many weeks in the future or past the outbreak peak was predicted to occur or to have occurred.

## Discussion

The main findings of our study demonstrate that given observation of new infected dengue cases in optimizing state variables and model parameters iteratively, the proposal of the combined SIR-EAKF model can timely return an accurate forecast, including forecast of unobserved states variable and adjustment of model parameters, for each week. Based on the returning estimator from forecast system, it can provide valuable reference for public health department of the government. For example, when a rapid uptrend of new infected dengue cases or infected mosquitoes is estimated, it warns the government to take necessary measures such as strengthening mosquito eradication to control the density of mosquitoes.

Regarding the seasonality of dengue fever, our study attaches importance to population fluctuation on mosquitoes and activity of dengue fever in explaining the seasonality of dengue pandemic ^32, 33^, which enhances the accuracy of peak time prediction to 87.5% with 6 or more consecutive weeks prior to the observed peak. In addition, the forecast of peak intensity and total incidence both have a good performance, leading accurate forecast of 75% and 62.5% one week ahead respectively, after an accurate peak time is captured. As a supplementary analysis, we also compare the combined SIR-EAKF model with GAM ^34, 35^. As shown in Supplementary Fig. 8, for each week, observation data range from the first week of 2011 to the current week, was used to fit GAM repeatedly, which can update the forecast of remaining period of dengue season forward. More details of GAM model construction and forecast strategy can be found in the appendix. In general, the prediction performance of the combined SIR-EAKF model is better than that of GAM, especially for the prediction of peak time which obviously can be predicted accurately earlier by the combined SIR-EAKF model.

However, it is inconsistent on forecast accuracy during high and low transmission season. Despite good performance on predicting the peak time for each situation, it is hard to forecast the low cases season which outbreak is rising from zero to peak in a short time, especially in seasons with peak intensity below 10. It may be caused by the greater impact of observation error on the fluctuation of reported dengue cases with increased dengue activity, or the excessively strict evaluation criteria of current low cases season. Nevertheless, the dengue prediction system has a good prediction for the season with high level of transmission observed, such as the 2014-2015 seasons. Moreover, for early warning of epidemic, it is more important to accurately predict the trajectory of high transmission season.

To simulate the actual transmission of dengue fever comprehensively, transmission path of mosquito-borne and direct transmission of mosquito are both considered in the dengue SIR model. However, some factors that may affect dengue transmission dynamics, such as the influence of environmental temperature ^36^ and humidity ^37^ as well as rainfall ^38^ on mosquito activities, different serotypes of dengue virus ^39^, imported cases ^32^, have not been fully considered. At present, limited by the available observation data, it is inappropriate to complicate the compartmental model into a high-dimensional model structure using only one observation data flow, which may be difficult to optimize. Only when more observed data flows are available, can more complex compartmental model be constructed to improve overall model performance.

Despite the settings of state variables and model parameters initialization for Guangzhou city, it can be widely adapted to various cities in southern China owning to the same characteristics of dengue outbreak as well as the flexibility of EAKF. As is known to all, dengue fever also spread from one city to another city (it can be seen from Supplementary Video S2), which reflects a faultiness of our study that the generated forecast doesn"t take account of the imported cases ^40-42^. As more data become available, we further introduce the combined SIR-EAKF model connecting each city by population flow to forecast the spatial diffusion of dengue ^20^.

In summary, a real-time forecast system for dengue fever constructed by assimilation observation and then iterative optimization model can well capture the trajectory of dengue for different scale of outbreak in Guangzhou. After familiar with the capabilities and limitations of the forecast, it brings lots of valuable reference to official on preventing outbreak of dengue likes that: on one hand, accurate prediction of peak time can help prevention measures well planned in advance; on the other hand, the power of prevention can measure by the peak intensity. Moreover, work like the weather forecast, can keep the public informed of the potential risk of dengue timely, and thus better supporting the whole society on prevention.

## Data Availability

All data included in this study are available upon request by contact with the corresponding author.

## Acknowledgements

We thank the staff members at the hospitals, local health departments, and county-, district- and prefecture-level Centers for Disease Control and Prevention for their great assistance in coordinating data collection.

## Supplementary Materials for

### Supplementary Methods

#### Dengue Model Development

As a mosquito-borne disease, dengue fever must be transmitted from person to person with the help of mosquitoes. Also the mosquito borne diseases are two-way, which can be transmitted from infected mosquitoes to susceptible humans, and from infected humans to susceptible mosquitoes ^1^. A compartmental model was constructed to describe the dynamics of dengue transmission by following equations:

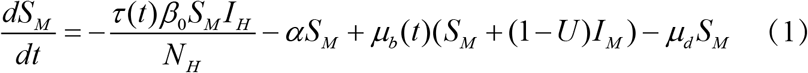

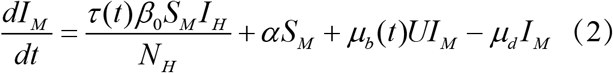

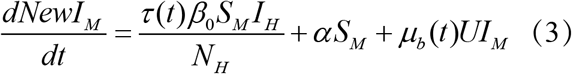

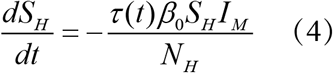

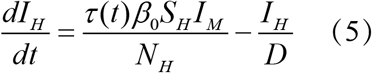

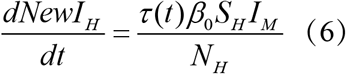

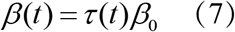

Where *S*_*M*_ is the number of susceptible mosquitoes, *I*_*M*_ is the number of infected mosquitoes, *NewI*_*M*_ is the number of new infected mosquitoes, *S*_*H*_ is the number of susceptible humans, *I*_*H*_ is the number of infected humans, *NewI*_*H*_ is the number of new infected human, *N*_*M*_ is the mosquito population, *N*_*H*_ is the human population, *α* is the rate of dengue seeding into the local model domain, *U* is the dissemination rate and is constant over an outbreak, *μ*_*b*_(*t*) is the mosquito birth rate at time *t, μ*_*d*_ is the mosquito death rate and is constant over an outbreak, *τ* (*t*) is the population transmission rate at time *t, β*_0_ is the basic contact rate between humans and mosquitoes, *β* (*t*) is the contact rate between humans and mosquitoes at time *t, D* is the mean infectious period for human.

This model uses a standard susceptible-infected-recovered (SIR) epidemiological construct in which all compartment are perfectly mixed. During each outbreak, population numbers of human are assumed as a constant for no birth or death was simulated while population numbers of mosquitoes are time-variant for the seasonal mosquito birth rate and constant death rate caused by ambient temperature ^2^.

In addition, dengue fever can be transmitted vertically through the mating of infected male mosquitoes with female mosquitoes ^1, 3^. Therefore, a constant parameter was introduced to represent the probability of vertical transmission of infected mosquitoes. Besides, the simulation was seeded with infected mosquitoes at a rate of one infected mosquito per 500,000 susceptible mosquitoes ^4^. The model assumed that all individuals met the following conditions: the susceptibility to the virus was the same; the infectivity of the virus was the same when infected; the mixed behavior related to the disease was the same.

#### Study Area

Guangdong province has always been one of the epidemic areas of dengue fever in China, especially its prefecture-level city Guangzhou. Guangzhou is the administrative center of Guangdong Province with a high degree of urbanization at an urbanization rate of 86%. Besides, there are more than 15.3 million permanent residents living in a total area of 7,434 square kilometers. The city occupies the central and southern part of Guangdong and adjacent to the Pacific Ocean in the South and near the Tropic of cancer with a subtropical climate all year round. The developed river system, agriculture and forestry through the city provide a ample breading sites for mosquitoes, the primary vectors of dengue fever.

#### Observed Human Dengue Cases

The individual surveillance data of dengue fever cases in Guangzhou from 2011 to 2018 were obtained from Guangdong CDC. All human dengue cases were diagnosed according to the diagnostic criteria for Dengue Fever (WS216-2008) enacted by Chinese Ministry of Health ^5, 6^. Among them, dengue virus subtypes were divided into four types, which were both included in the statistics. Also, an index of whether they were imported or local cases were recorded and only the local cases were included in this study to avoid the uncertainty of import cases. Dengue cases were aggregated by week according to the date of illness onset with each week defined as Sunday to Saturday. Fig. 1 b-h shows the Weekly observations of dengue cases from Jun 2011 to Dec 2017 in Guangzhou. This study assumed that the error variance of observation was associated with each weekly observation. The observational error variance (OEV) was given by the empirical rule.

#### Mosquito information

The surveillance data of mosquito density in Guangzhou from 2011 to 2018 were obtained from Guangdong CDC. There are two index of mosquito larval density was considered including the Breteau Index (BI) and the Mosquito Oviposition Index (MOI). The Breteau index is defined as the number of positive containers per 100 households inspected. Besides, the mosquito oviposition index is defined as number of ovitrap with positive adult and egg of Aedes albopictus / number of effective ovitrap^7^.

To construct the *μ*_*b*_(*t*) of compartmental model, MOI information was converted into mosquito natural birth rate by following procedure. First, Fourier transform method was used to smooth the MOI. In a dengue season, the standardized natural growth proportion sequence was obtained by dividing each MOI by the MOI where the first derivative was the minimum. We assumed the oviposition period of mosquitoes is 16 days ^8^, so natural birth rate was calculated as the standardized natural growth proportion sequence, which is rooted 16 power and then minus 1. Finally, the *μ*_*b*_(*t*) can be calculated as the natural birth rate minus the constant mosquito death rate *μ*_*d*_.

Assuming that the residuals of natural birth rate in each time point of several year were normally distributed, the natural birth rate time series of each ensemble were randomly select from the distribution which the mean and standard deviation were calculated by historical data from 2011 to 2018.

#### Meteorological data

Daily meteorological data on ambient temperature, maximum temperature and rain volume for the same period were publicly available from the China Meteorological Data Sharing System (http://data.cma.cn/). Several studies have shown the association between the reproduction of dengue virus in mosquitoes and the ambient temperature ^9, 10^. Based on other study, a function was constructed to represent the relationship between ambient temperature and population transmission rate which is the transmission probability of dengue virus by mosquito bites ^9^. The function is list as follows:

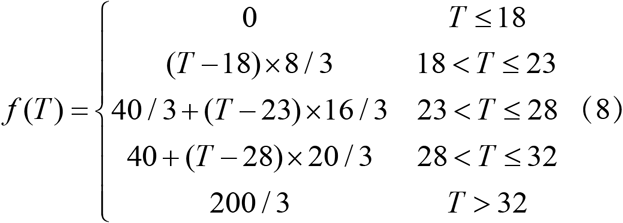

where *T* is the ambient temperature. *f* (*T*) is the translated population transmission rate. Then the time series of population transmission rate can be confirmed.

Assuming that the residuals of population transmission rate in each time point of several year were normally distributed, the population transmission rate time series of each ensemble were randomly select from the distribution which the mean and standard deviation were calculated by historical data from 2011 to 2018.

#### Runge-Kutta method for SIR model

In order to improve the accuracy of forward prediction of SIR model, the classical Runge-Kutta method was used in the iteration of state in SIR model. When estimating the slope *k* of *x*_*i*−1_ and *x*_*i*_, it is obviously inaccurate to use only the slope *k*_1_ of *x*_*i*−1_, which will seriously increase the error. However, Runge-Kutta method is to estimate the slope values *k*_1_, *k*_2_,…, *k*_*m*_ of several points on [*x*_*i*−1_, *x*_*i*_] and the weighted average of them is taken as the average slop of *y*(*x*) on [*x*_*i*−1_, *x*_*i*_]. Due to restrain the error by Runge-Kutta method, it can solve the iterative process of state variables more accurately when applied to the above SIR model. The classical Runge-Kutta algorithm used in this study is as follows:

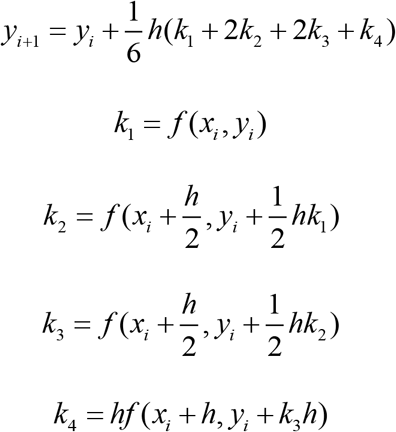

where *k*_1_ is the slope at the beginning of the time period; *k*_2_ is the slope at the zmidpoint of the time period, and the slope *k*_1_ is used to determine the value of y at the point 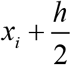 by Euler method; *k* is the slope at the midpoint of the time period, but the slope *k*_2_ is used determine the value of y; *k*_4_ is the slope at the end of the time period, and its y value is determined by the slope *k*_3_.

#### Filtering Method of EAKF

The ensemble adjustment Kalman filter (EAKF) is a kind of data assimilation technology which can estimate the true state given observations and simulation of that state. The algorithm of EAKF is introduced below.

Kalman filter, in general, assume that the observations 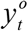 at time *t* are allowed to be divided into several uncorrelated subsets 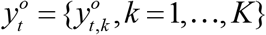. A joint state-observation vector for a given *t* and *k* can be written as *z*_*t,k*_ = [*X*_*t*_, *h*_*t,k*_ (*X, t*)], where *h*_*t,k*_ (*X, t*) is an observation operator and *X*_*t*_ represents the state of model system at time *t*. The length of joint state vector is *n* + *m*, where *n* is the size of *X*_*t*_ and *m* is the size of the observational subset 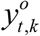. The estimated observation values can be calculated by equation *y*_*t, k*_ = *Hz*_*t, k*_, where *H*_*i, j*_ = 1 for *j* = *i* + *n,i* = 1,K, *m* and *H*_*i, j*_ = 0 for all other elements. The conditional distribution after making use of the next subset of the joint state vector is

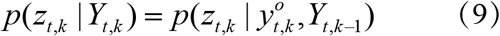

Where *Y*_*t, k*_ represents an superset of *k* observation subsets before time *t* +1. Returning to the approach of Jazwinski, Bayes"s rule gives

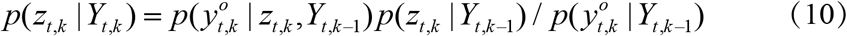

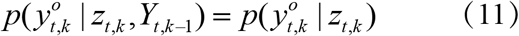

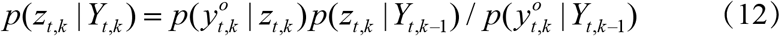

Equation (12) shows new sets of observations modify the prior joint state conditional probability distribution through the predictions based on previous observation sets.

Assuming that the observation error and prior distribution satisfy the Gaussian distribution, then the equation (12) can be represented by the convolution of two Gaussian distributions, and the distribution of result is still Gaussian distribution. So the covariance and mean of *p*(*z*_*t, k*_ | *Y*_*t, k*_) can be written as

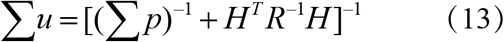

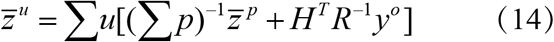

where *R* is the error covariance 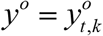. A linear operator *A* is of observation introduced to get the updated ensemble.

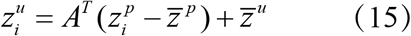

EAKF adjusts ensemble of model simulation state variables to true state. Using cross ensemble co-variability, unobserved state variables and parameters are also updated. For further details of EAKF algorithm see Anderson ^11^.

#### Description of SIR-EAKF system

Previous studies ^12, 13, 14^ have been used the EAKF assimilation method conjunction with a variety of compartmental epidemiological model to assimilate the observation data and update the simulation data based on Bayes" rule, which also shows a better performance than other filtering methods ^15, 16^. In this study, a 300-member ensemble simulation of the SIR compartmental model (Equations 1-7) was run in conjunction with the Guangzhou dengue cases data and the EAKF. There were 6 state variables and 3 parameters *X*_*t*_ = (*S*_*M*_, *I*_*M*_, *S*_*H*_, *I*_*H*_, *D, β, N, NewI*_*M*_, *NewI*_*H*_) and weekly observations of human dengue cases 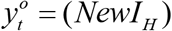 included in the filtering frame work to estimate the true state and parameter of system.

The model assumed that the mosquito population size was time-varying for the combine affection of the seasonal birth rate and constant death rate. Besides, the scaling of observed human dengue cases to the model simulation was assumed to be one. Whenever an observation was available, the observation was reported once a week in this study. The ensemble dengue SIR model were used to predict the next state variables and then updated with EAKF on observation state variables. In addition, the inter-variable relationships were assumed to be linear. Consequently, a cross ensemble co-variability method was used to adjust the unobserved state variables and parameter (*S*_*M*_, *I*_*M*_, *S*_*H*_, *I*_*H*_, *D, β, N, NewI*_*M*_) by multiplying the ensemble covariance with the observation adjustments in EAKF algorithm. The SIR-EAKF model was then integrated forward to the next observation using the update (posterior) state variables and parameters and the data assimilation updating process is repeated. Over time, the observed dengue cases were used for recursive adjustment to optimize the model state variables and parameters, so that the integrated model simulation can better simulate the local outbreak dynamics.

#### Self-determination method for the epidemic affected population size and combination with SIR-EAKF (combined SIR-EAKF)

Due to the huge difference between peak intensity of each year, it is too big a fixed epidemic affected population size given for the season with a small peak incidence to predict, and too small a fixed epidemic affected population size given for the season with a big peak incidence to predict. Consequently, it is necessary to consider a suitable epidemic affected population size, which can take into account seasons with difference peak incidence.

The epidemic affected population size is obviously related to the spatial distribution of the number of new infected dengue cases. As shown in Supplementary Fig. 9 A-G, the Guangzhou city was subdivided into a number of 25 kilometers of hexagon. Besides, the area of hexagon with color indicated the epidemic area of the corresponding year (the area must have 1 or larger than 1 dengue cases). The onset week of hexagon area was distinguished by color. It can be seen in the figure that there were a same spatial distribution form of spreading outward with a certain central point, which was in line with the spatial transmission characteristics of the epidemic. The figures in set of A-G were the distribution of epidemic affected population size (blue) and its area size (red) in the area of hexagon with dengue cases. Assuming that the population was evenly distributed in space, the epidemic affected population size was calculated by the number of permanent residents in each county, which were obtained from statistical yearbook. These figures were indicated the epidemic affected population size and area size both had a pattern seen like the time series of new infected dengue cases which had a peak value in October.

Supplementary Fig. 9 H shows a scatter plot of the logarithm of the number of new infected dengue cases + 1 and the epidemic affected population size and area size in each month from 2011 to 2017. Different color was used to distinguish each year, and the distance to the peak time by month of corresponding year was used to distinguish by the size of the points. It can be seen that there are a larger epidemic affected population size and area size in each year when getting closer to the peak time. Furthermore, we gave a Pearson correlation analysis of the epidemic affected population size and area size with log(new infected dengue cases + 1), which both gives a statistical significance high Pearson correlation coefficient.

In this study, in order to solute the problem of different scale of outbreak in each epidemic season, we suggest to use a simple self-determination method for the epidemic affected population size. The epidemic affected population size was calculated from the historical data and used as a fixed parameter of SIR-EAKF. It should rerun the SIR-EAKF model to assimilate observed data when the epidemic affected population size was update. In particular, first of all, the average proportion of weekly new infected dengue cases in the total number of infected dengue cases in that year was calculated according to the history data from 2011 to 2017 (except year of 2014 which have an abnormally high value). After that, the epidemic affected population size in current time was estimated through the average proportion sequence and the current observation. By estimating and updating the epidemic affected population size (the fixed parameter of SIR-EAKF) at time *t*, the assimilation and update process was need to rerun the time period of {1, 2,…,*t*}. Moreover, in the combined model, this method was set to run from the fifth week to the average peak time of 22 weeks to reduce the amount of computation, and the lower limit of the epidemic affected population size was 300 and the upper limit was 1,500,000.

#### Generation of Synthetic Truth and Observation

We generated a synthetic model-simulated dengue season to validate the optimization EAKF of dengue SIR model combined with Self-determination method for the epidemic affected population size. The synthetic dengue season was generated by free simulation of dengue SIR model (Equation 1-6) and defined as the “true”. The simulation was initiated with *N*_*M*_ (0) = *N*_*H*_ = 12000 / 0.7, *S*_*H*_ (0) = 0.8*N*_*H*_, *I*_*H*_ (0) = 0, *S*_*M*_ (0) = *N*_*M*_ − *I*_*M*_ (0), *I*_*M*_ (0) = 1.2, *D* = 6, *β*_0_ = 0.05, *U* = 0.25, *μ*_*d*_ = 1/ 15, *α* =1/ 500000. The simulation generated by this parameter combination make a good representation of the mean reported weekly dengue cases for Guangzhou city from 2011 to 2017 except 2014. Synthetic observations of dengue cases were then generated by adding normally distributed random observational error (mean 0 and standard deviation equal to the observation * 0.025) to the truth. These synthetic error-laden observational records reported dengue cases were then used for assimilation in the combined SIR-EAKF system.

#### Application of Synthetic Observations to the Model-Inference System

Using the synthetic observations of weekly dengue cases and its defined OEV, the combined SIR-EAKF framework could make a result whether the forecast were appropriately estimate and adjust all state variables and model parameters. In the combined SIR-EAKF system, 300 ensembles was set and used to simulate. The state parameters of each ensemble member was initialized as: *S*_*M*_ (0) = *N*_*M*_ (0) − *I*_*M*_ (0), *I*_*M*_ (0) = *U* (0, *N*_*M*_ /1000), *S*_*H*_ (0) = *N*_*H*_ ×*U* (0.7, 0.9) − *I*_*H*_ (0), *I*_*H*_ (0) = *U* (0,1); and model parameters were randomly selected from uniform distributions: *D* = *U* (5, 7), *β*_0_ = *U* (0.045, 0.055), *N* = *N*_*H*_ = *N*_*M*_ (0) = *N*_*p*_ / *U* (0.6, 0.8), and average of peak time was set as the 22th week of the dengue season. The simulation was seeded with infected mosquitoes, *α*, during integration over season at a rate of 1 in 500,000. In addition, each ensemble member with the constant model parameter: *U* = 0.25 and *μ*_*d*_ = 1/ 15 over an outbreak. For each outbreak, model optimization began from the 20th week of current year to the 19th week of next year. The dengue SIR model was run by day and the assimilation of synthetic observations of dengue cases was run by week using EAKF.

Over all, the ensemble posterior mean state variable and parameter estimates were well constrained (Supplementary Fig. 1). Affected by the self-determination method of epidemic affected population size, the estimation of susceptible mosquitoes *S*_*M*_ and susceptible human *S*_*H*_ is lower than the true in the initial stage of the dengue season for a low epidemic affected population size *N*. As time progresses and more information is observed about the outbreak, the unobserved state variables *N, S*_*M*_ and *S*_*H*_ were well captured. Besides, the observed state variable, new infected human dengue cases *NewI*_*H*_, and the unobserved state variables, new infected mosquitoes dengue cases *NewI*_*M*_, infected human dengue cases *I*_*H*_, infected mosquitoes dengue cases *I*_*M*_, were also well captured over the dengue season. In addition, the combined model also estimated the epidemiological parameters which were helpful to describe the epidemic characteristics of infectious diseases. Specifically, the infectious period for human, *D*, fluctuates around the true owning to the change of susceptible population. Due to the lack of initial susceptible population, model parameters *D* were adjusted higher than the true in response to the correction of observed variable. When an appropriate epidemic affected population size was given in a re-run SIR-EAKF, the susceptible population was captured and the parameters *D* were slightly adjusted closer to the true. It was the same reason why the basic contact rate between humans and mosquitoes, true. *β*_0_, fluctuates around the

#### Forecast Procedure

Weekly ensemble forecasts of future dengue cases are obtained after assimilating new observation data. At the start of a particular dengue season, given a suitable set of initial conditions can help compartment model grasp the trend of outbreak and reduce the dependence on EAKF. In practice, the state variables and model parameters are optimized by repeatedly adjusting the compartment model with iterative simulation and assimilation of observation. The state variables and model parameter can be aligned to the true in this way, so as to make a more accuracy prediction to the outbreak. Using the latest posterior estimates of the state variable and model parameters, the compartmental model (Equations 1-6) is integrated to generate those forecasts until the end of outbreak. In addition, during the 5th week to the 22nd week of dengue season, the SIR-EAKF system is rerun with a update epidemic affected population size.

#### Retrospective Forecast

The forecast procedure was used to generate retrospective forecasts of dengue outbreaks from 2011 to 2017. An ensemble compartmental-model was initiated with a 300-member ensemble for each outbreak season (the 20th week of current year to the 19th week of next year). Each ensemble member state variables was initialized with: *S*_*M*_ (0) = *N*_*M*_ (0) − *I*_*M*_ (0), *I*_*M*_ (0) = *U* (0, *N*_*M*_ /1000), *S*_*H*_ (0) = *N*_*H*_ ×*U* (0.7, 0.9) − *I*_*H*_ (0), *I*_*H*_ (0) = *U* (0,1) ; and model parameters were randomly selected from uniform distributions: *D* = *U* (5, 7), *β*_0_ = *U* (0.045, 0.055), *N* = *N*_*H*_ = *N*_*M*_ (0) = *N*_*p*_ / *U* (0.6, 0.8), and average of peak time was set as the 22th week of the dengue season. The simulation was seeded with infected mosquitoes, *α*, during integration over season at a rate of 1 in 500,000. In addition, each ensemble member with the constant model parameter: *U* = 0.25 and *μ*_*d*_ = 1/ 15 over an outbreak. For each outbreak, model optimization began from the 20th week of current year to the 19th week of next year. As an example, the retrospective forecast for 2013-2014 dengue epidemic seasons can be seen in Supplementary Video S1.

#### Comparison of Prediction Accuracy between GAM and the combined SIR-EAKF

Previous studies have been used the GAM to make a prediction of the outbreak of dengue fever with a well performance^17, 18^. We had compared the prediction accuracy of GAM and SIR-EAKF using three prediction accuracy indexes (peak time, peak incidence, and total incidence) for the 2013-2014 through 2016-2017 dengue seasons. Here, we introduced the construction of GAM model and the prediction strategy. Imitating Xu Lei"s study, the GAM model was established in two steps. First of all, we contribute the GAM model of the mosquito density and the dengue incidence as follows:

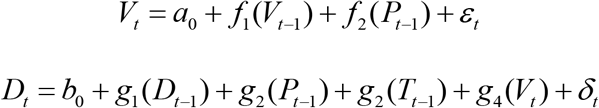

where *V*_*t*_ and *D*_*t*_ represent the weekly mosquito density and dengue incidence in week *t* respectively, *T*_*t* −1_ and *P*_*t*−1_ represent the weekly average of highest temperature and the number of days with rainfall for week *t* −1 respectively, the functions *f*_1_, *f*_2_ are linear functions, the functions *g*_1_, *g*_2_, *g*_3_, *g*_4_ are smooth functions of natural cubic spline. Using the prediction of *V*_*t*_ from GAM of mosquito density, the GAM of dengue incidence then predicted the dengue incidence *D*_*t*_ in *t* week.

However, the above GAM model can only forecast one week ahead and unable to forecast the rest of dengue season. Consequently, the above GAM model was modified according to the number of lead weeks (*n*) as follows:

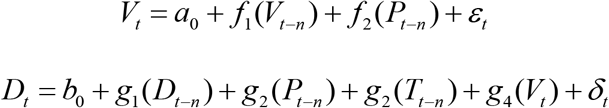

Although the above the steps can help GAM model predict several weeks ahead, it also bring a huge amount of computation. Through the prediction strategy of GAM, it can predict the dengue incidence of *t,t* +1,…,53 at week *t*, which was used to compare with the corresponding prior forecast of SIR-EAKF model. To compare with the SIR-EAKF model, the similar posterior forecast is given by the fitting value of GAM model at week *t*. In this way, the GAM model can predict the whole epidemic season at each week, which was similar to the SIR-EAKF framework. Then, it is available to compare the forecast accuracy of GAM model and SIR-EAKF model with peak time, peak incidence and total incidence.

#### Analysis of Retrospective Forecasts

The quality of the retrospective seasonal forecasts was analyzed through comparison to observations to determine how well each ensemble forecast estimated the peak timing and peak magnitude and the total magnitude of the seasonal number of dengue cases. For all 3 metrics we compared the ensemble mean trajectory with observed outcomes. Forecasts were considered accurate if: 1) it peaked within ±1 week of the observed peak of new infected dengue cases; 2) the maximum new infected dengue cases was within ±25% or ±1 of the observed peak incidence; 3) the total number of new infected dengue cases over the entire season was within ±25% or ±1 of the observed, whichever was larger. As an additional analysis, forecasts were grouped by prediction lead, e.g., how many weeks in the future or past the outbreak peak was predicted to occur or to have occurred. All forecasts with the same lead were grouped and the fraction of accurate forecasts was quantified.

---

**Supplementary Figure 1.**
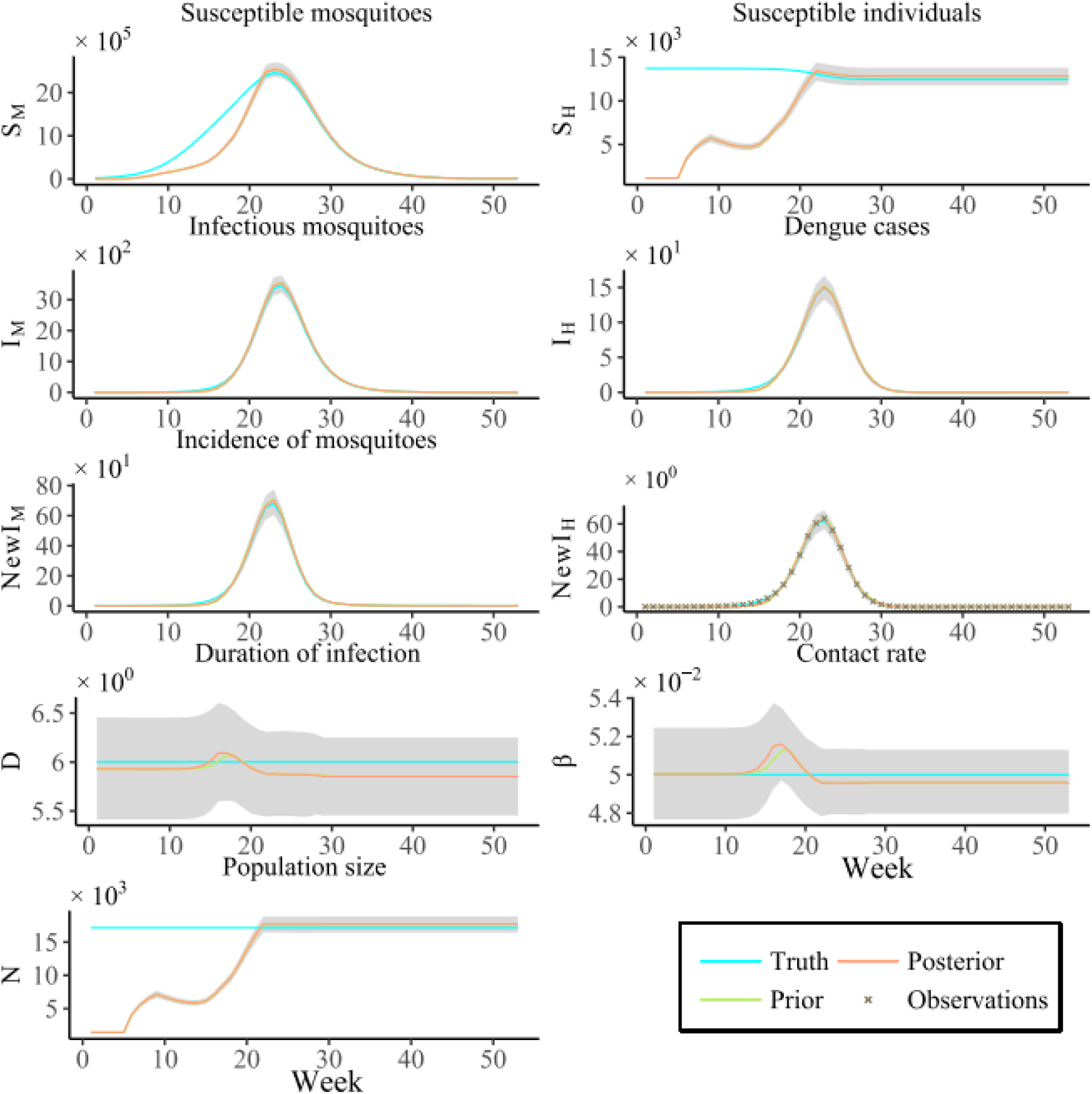
Inference of state variables and parameters in the SIR-EAKF model. Prior (green) and posterior (red) mean estimates of state variables *S*_*M*_, *S*_*H*_, *I*_*M*_, *I*_*H*_ and parameters *D, β*, and *N* as inferred by SIR-EAKF model, were displayed. The grey area is the spread of the ensemble forecast between the 25th and 75th percentile. The synthetic outbreak (blue) was constructed by free simulation with the mean of weekly new infected for the 2011-2012 through 2017-2018 seasons except 2014-2015 season. (*D* = 6, *β*= 0.5 and *N* = 12,000). The observation (cross) was computed by adding disturbance from synthetic true.

**Supplementary Figure 2.**
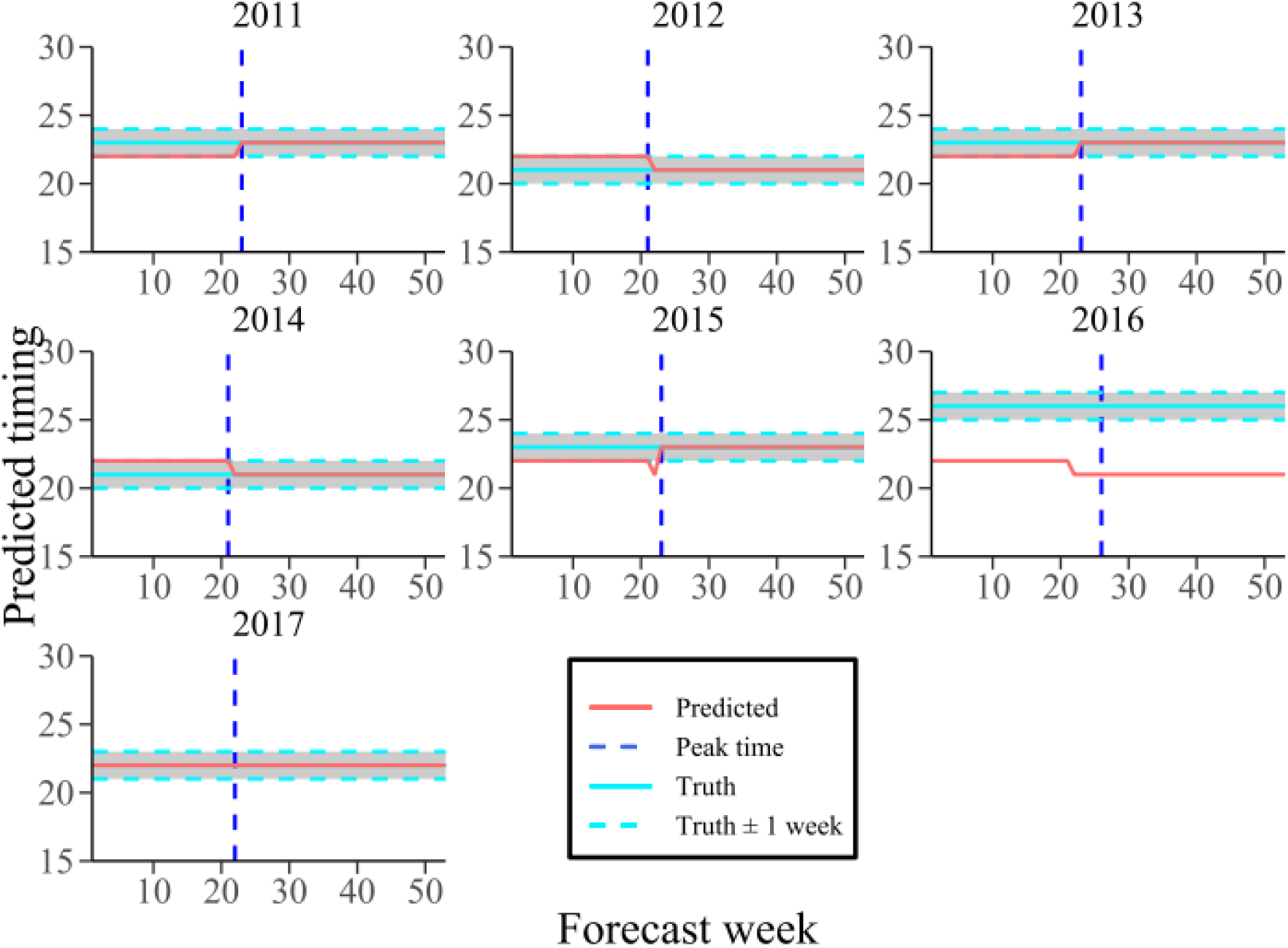
Weekly SIR-EAKF forecasts of peak timing for the 2011-2012 through 2017-2018 seasons. The true timing of peak intensity occurs in the season (horizontal light blue solid line) and its accuracy interval (horizontal light blue dotted line; ± 1 week of the observed) and the observed peak (vertical royal blue dotted line) were also shown. Note that SIR-EAKF forecasts (red) to the left of vertical line were made prior to the peak and forecasts to the right were made after the true peak had passed.

**Supplementary Figure 3.**
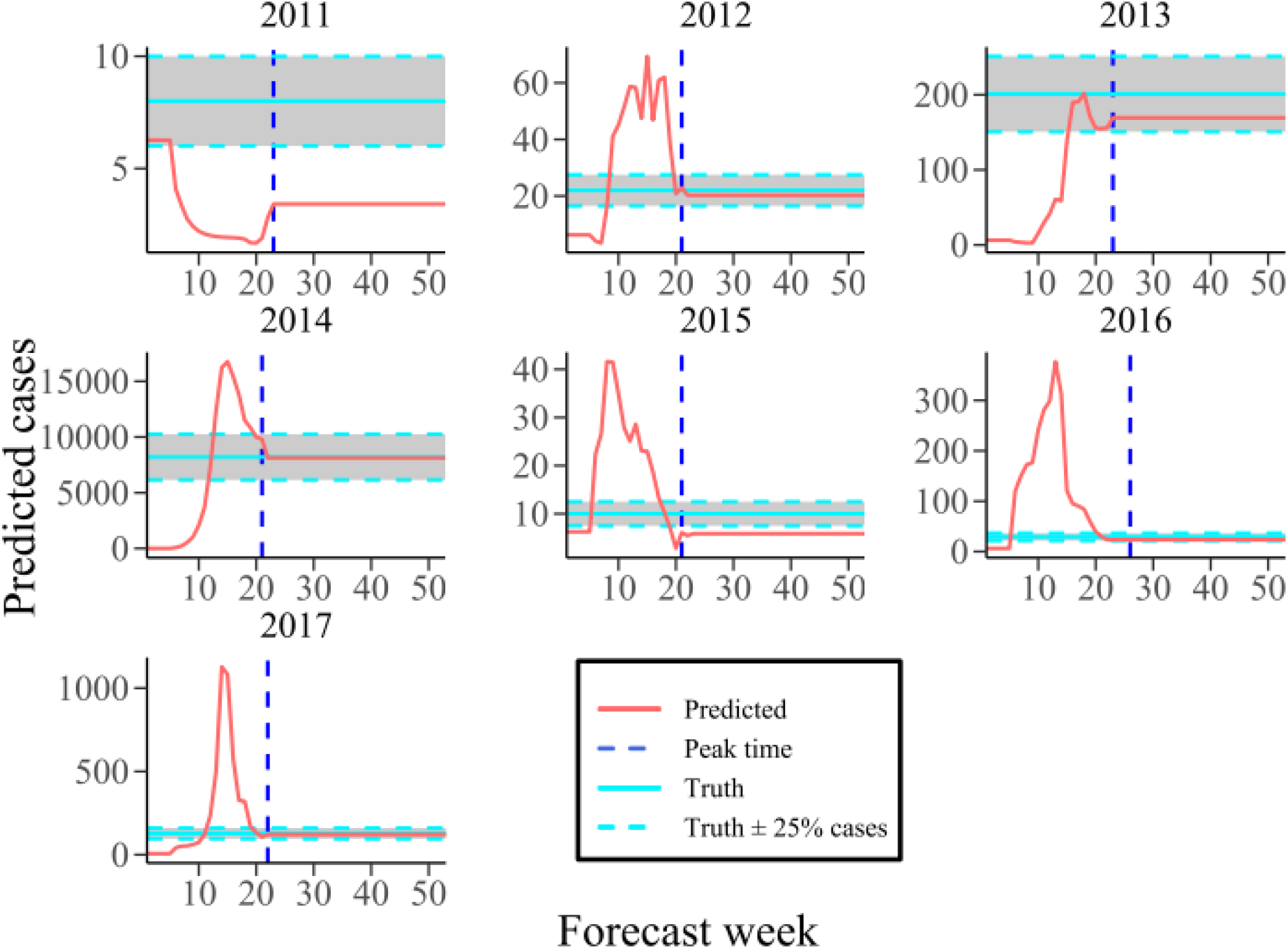
Weekly SIR-EAKF forecasts of peak intensity for the 2011-2012 through 2017-2018 seasons. The true peak intensity of the season (horizontal light blue solid line) and its accuracy interval (horizontal light blue dotted line; ± 25% or ± 1 cases of the observed) and the observed peak (vertical royal blue dotted line) were also shown. Note that SIR-EAKF forecasts (red) to the left of vertical line were made prior to the peak and forecasts to the right were made after the true peak had passed.

**Supplementary Figure 4.**
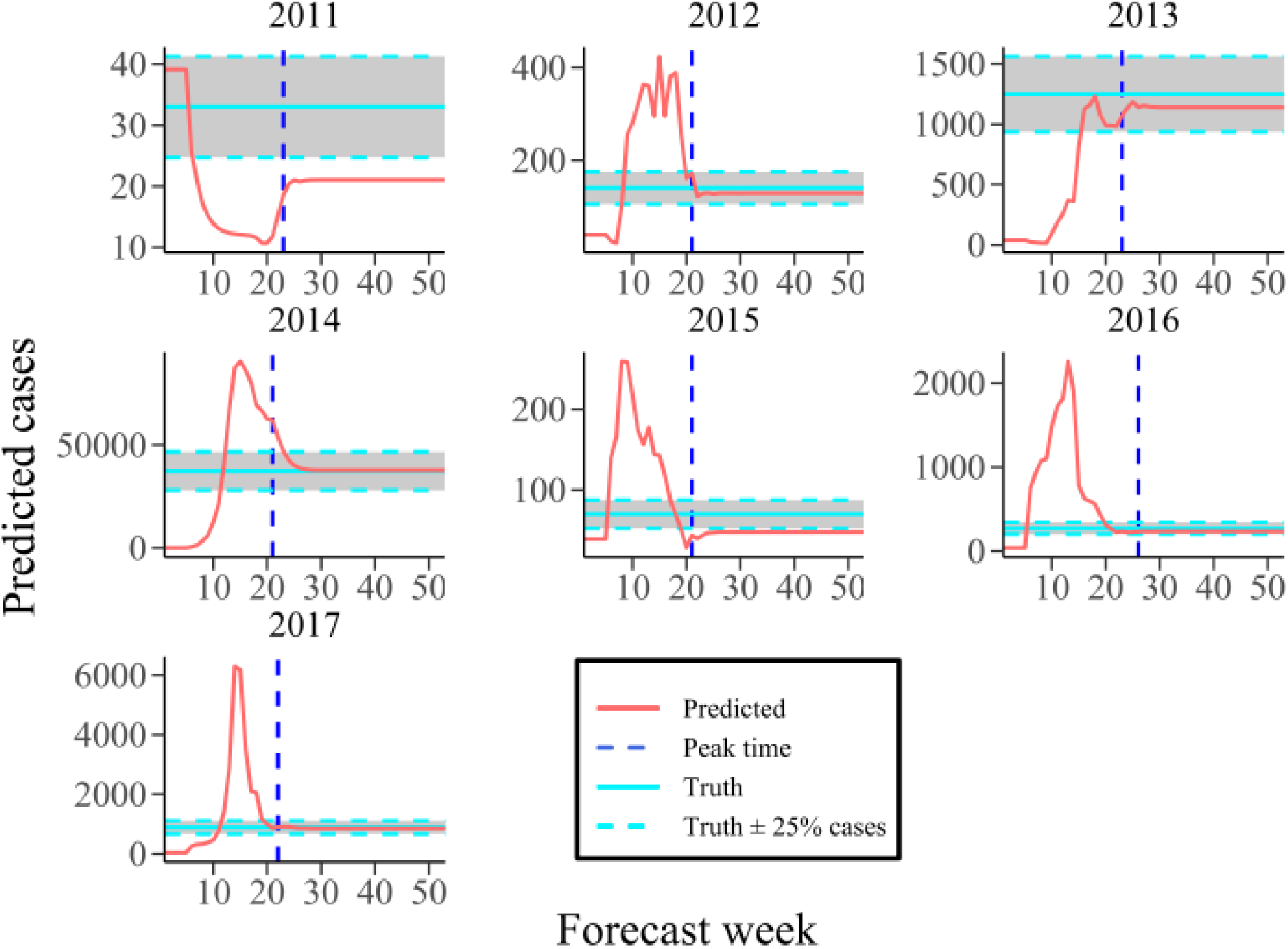
Weekly SIR-EAKF forecasts of total incidence for the 2011-2012 through 2017-2018 seasons. The true total incidence of the whole season (horizontal light blue solid line) and its accuracy interval (horizontal light blue dotted line; ± 25% cases of the observed) and the observed peak (vertical royal blue dotted line) were also shown. Note that SIR-EAKF forecasts (red) to the left of vertical line were made prior to the peak and forecasts to the right were made after the true peak had passed.

**Supplementary Figure 5.**
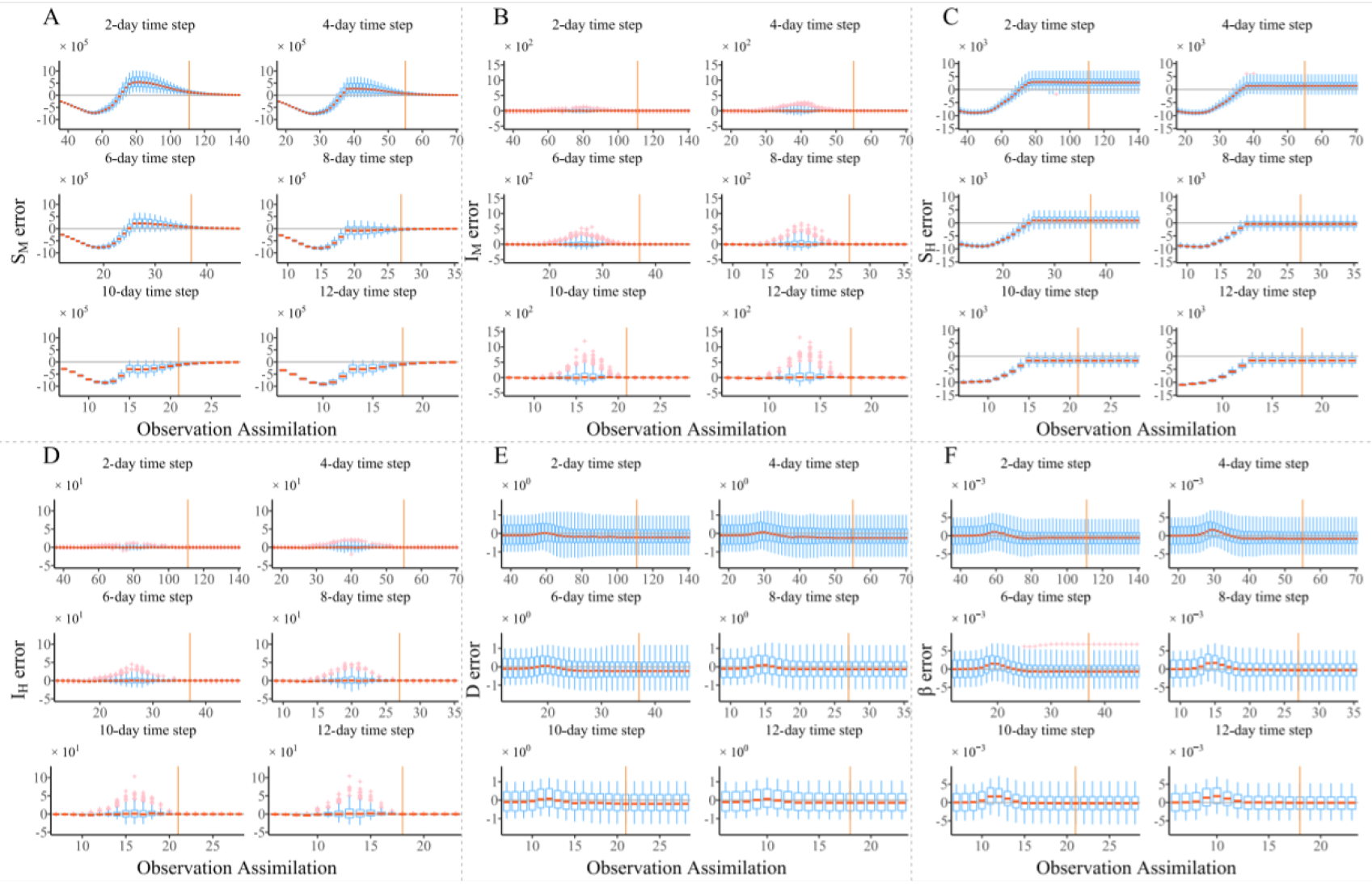
Results of SIR-EAKF assimilation sensitivity tests for changes to the time interval between observations. **a** Time series of the distributions of mean ensemble mosquito susceptible error relative to the synthetic truth for observations made every 2, 4, 6, 8, 10, and 12 days. 300-member EAKF assimilation runs were performed to generate each suplot. The box and whisker form shows the distribution of ensemble posterior mean error following each observation assimilation, including error median (red segment), 25th and 75th percentiles (blue box), extremes (whiskers), and outliers (pink cross) following each observation assimilation. For clarity, the box and whisker distributions are shown for every other assimilation for the 2-d time-step interval (*Top Left*). **b** Same as **a** but for mean ensemble new infected dengue cases. **c** Same as **a** but for mean ensemble human susceptible. **d** Same as **a** but for mean ensemble new infected mosquitoes. **e** Same as **a** but for the parameter *D* (mean infectious period). **f** Same as **a** but for the parameter *β* (contact rate).

**Supplementary Figure 6.**
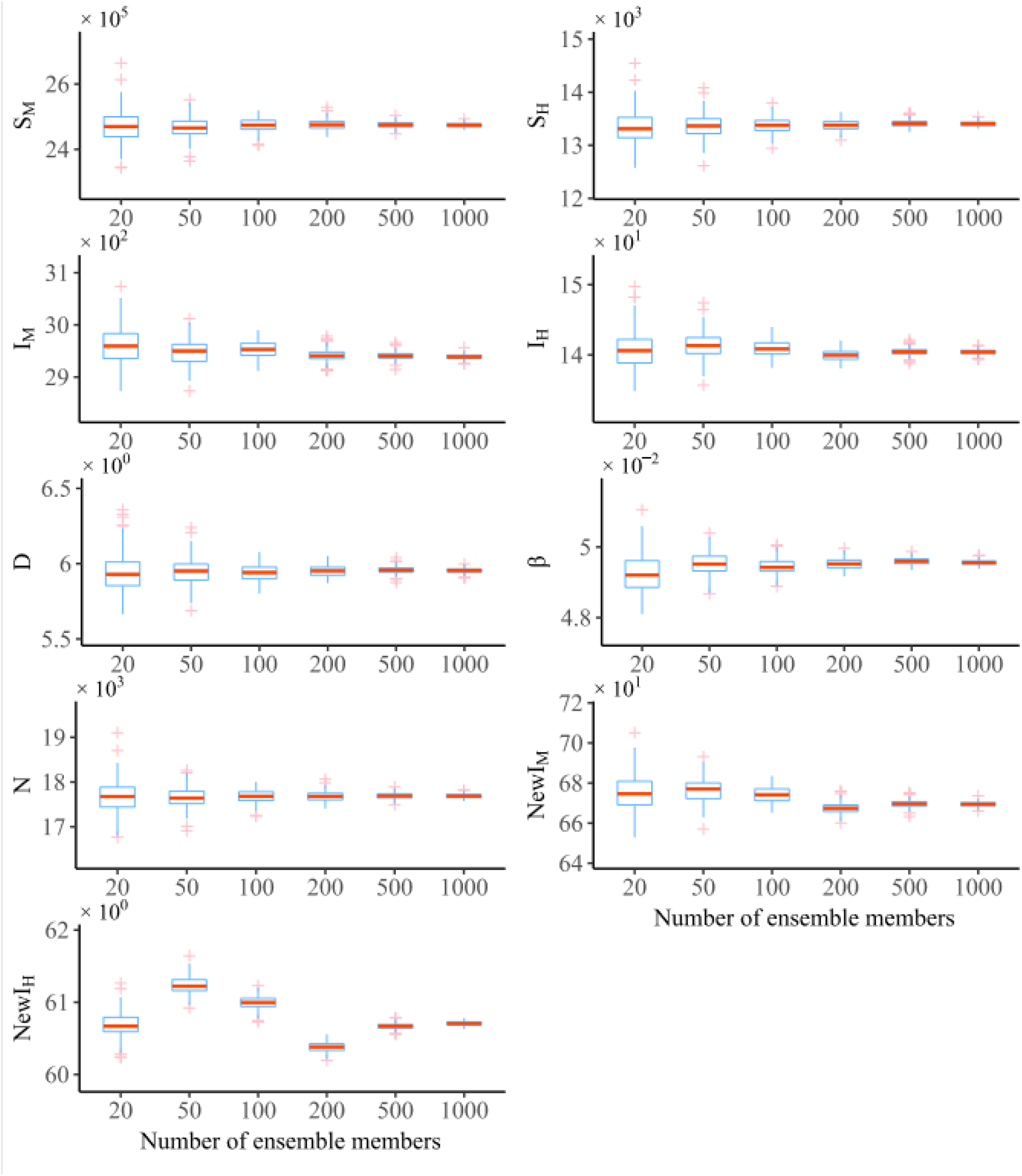
Results of SIR-EAKF assimilation sensitivity tests for different ensemble sizes. Ensemble sizes tested are 20, 50, 100, 200, 500, and 1,000 members. The distribution of mean ensemble estimator for 250 EAKF assimilation runs for each of these ensemble sizes is shown at week 22 for *S*_*M*_, *S*_*H*_, *I*_*M*_, *I*_*H*_, *D, β, N, NewI*_*M*_, *and NewI*_*H*_.

**Supplementary Figure 7.**
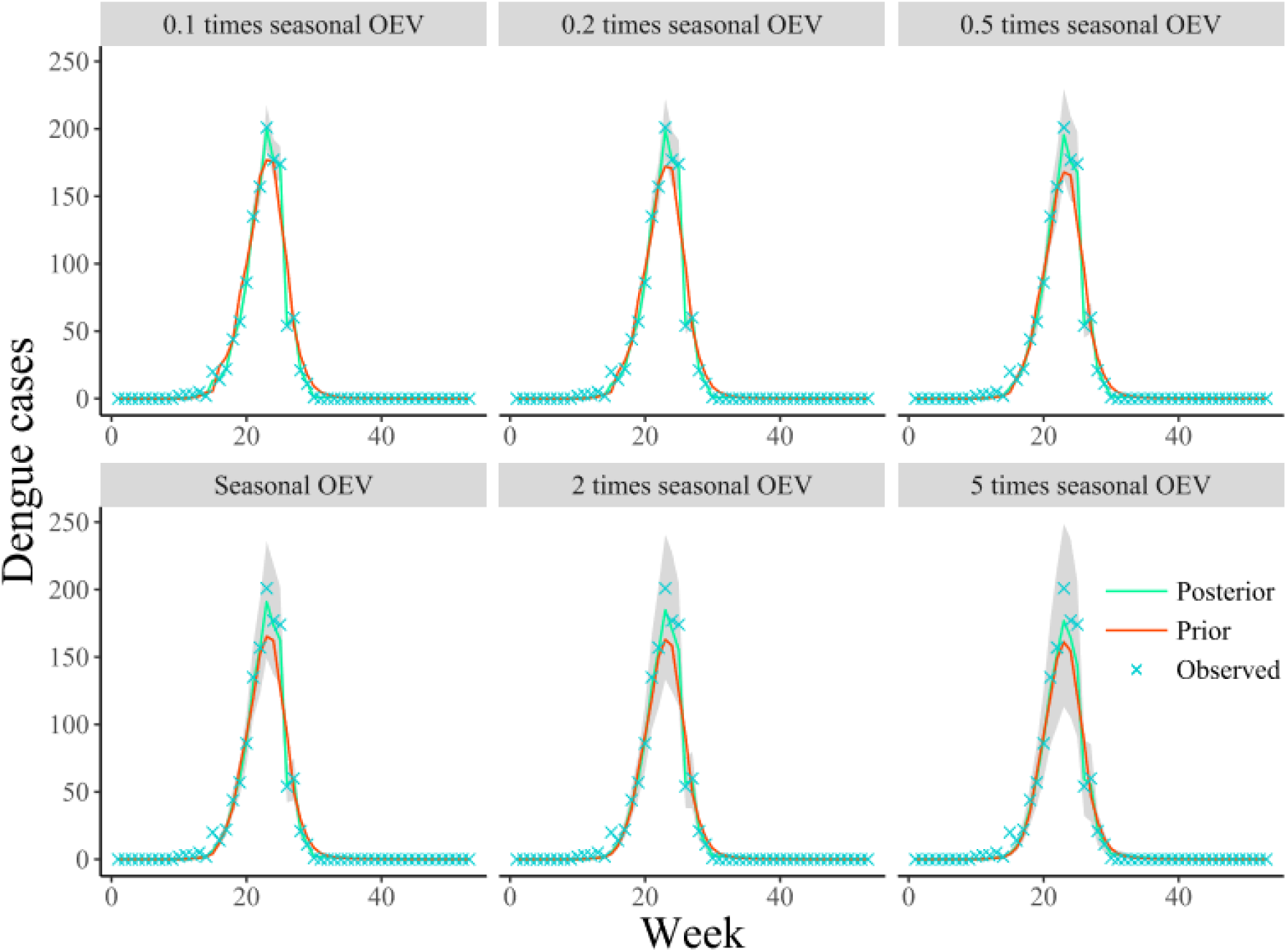
Results of sensitivity tests using different seasonal, time-varying OEV with weekly observed dengue cases for 2013-2014 dengue season. The seasonal OEV is scaled by a multiple of 0.1, 0.2, 0.5, 1, 2, or 5. All runs use a 300-member ensemble and 7-d interval between observations. Each subplot shows the prior (red) and posterior (green) mean ensemble new infected dengue cases with different scaling of 250 EAKF assimilation along with observations (cross). Also, the mean spread of the ensemble forecast between the 25th and 75th percentile are shown in grey area.

**Supplementary Figure 8.**
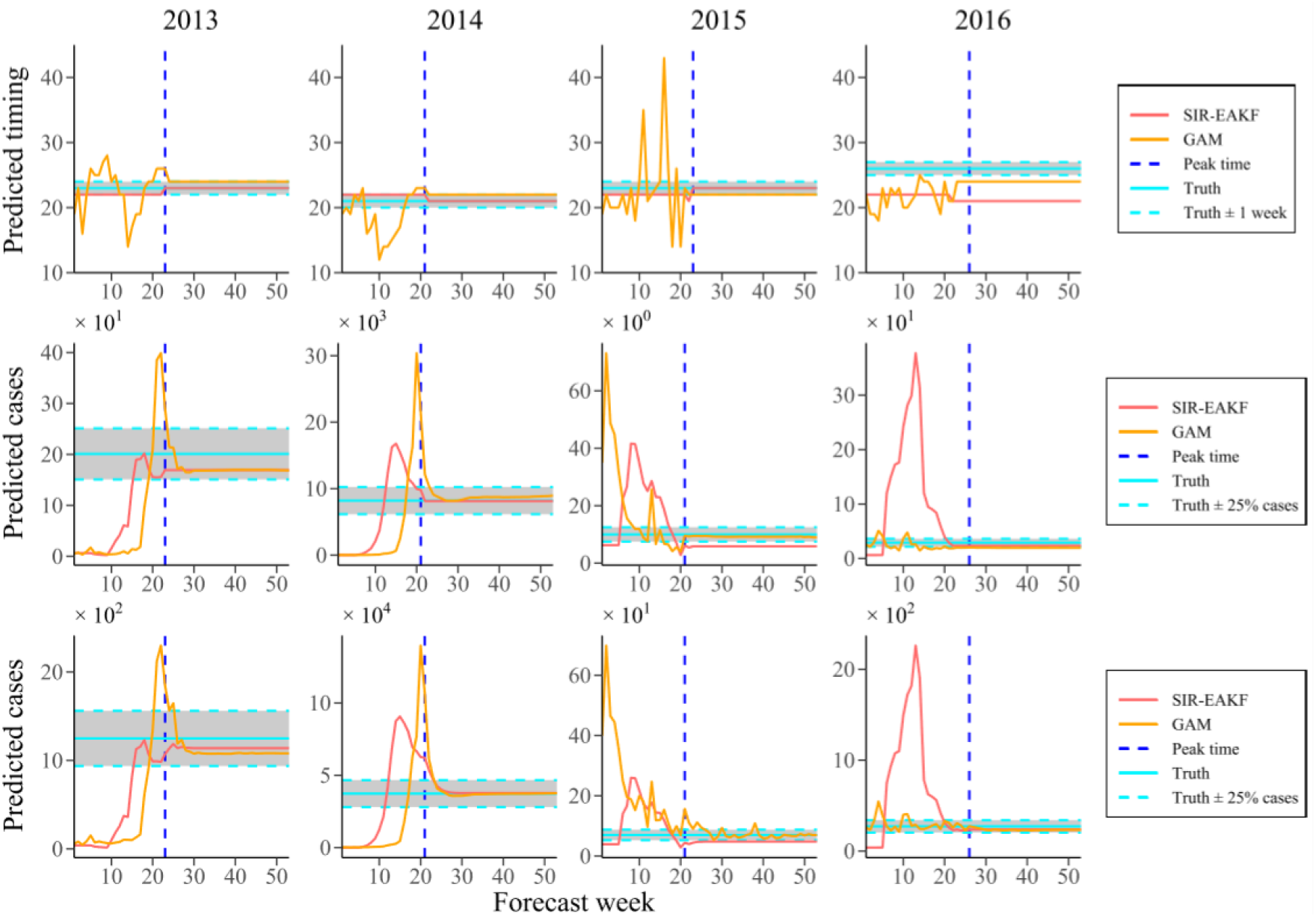
Weekly SIR-EAKF and GAM forecasts of predicted index (peak timing, peak intensity and total incidence) for the 2013-2014 through 2016-2017 seasons. The peak timing, peak intensity and total incidence are shown in sequence from top to bottom. The true index (horizontal light blue solid line) and its accuracy interval (horizontal light blue dotted line; ± 1 week / ± 25% cases of the observed) and the observed peak (vertical royal blue dotted line) were also shown. Note that SIR-EAKF forecasts (red) and GAM forecasts (golden) to the left of vertical line were made prior to the peak and forecasts to the right were made after the true peak had passed.

**Supplementary Figure 9.**
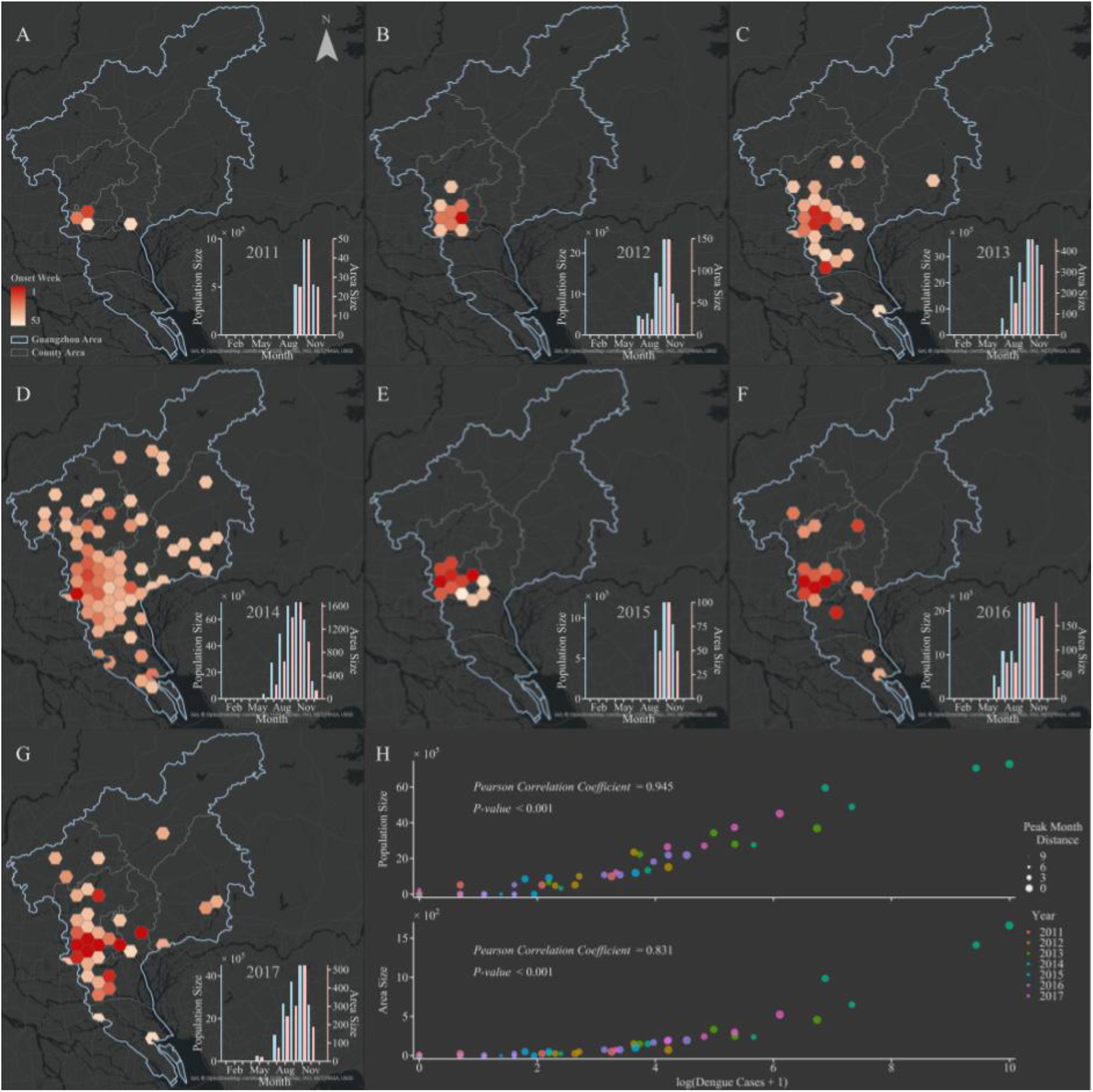
The spatial transmission pattern of dengue fever and its monthly changes of population size and area size affected by the epidemic from 2011 to 2017. **A-G** The temporal and spatial spread of the epidemic in Guangzhou from 2011 to 2017. The onset week of administrative regions has shown. Small figures inset of A-G show the monthly change trend of population size and area size affected by the epidemic. **H** Scatter plot of the relationship between monthly new infections and the population size (area size) affected by the epidemic. The distance of peak month and different years are respectively distinguished by the size and color of scatter. And the Pearson correlation test are also shown.

**Supplementary Video S1**.

**Animation of 300-member SIR-EAKF retrospective forecasting for 2013-2014 season**. Ten weekly forecast of dengue cases for 2013-2014 season. The purple lines are the ensemble mean forecasts, the grey area is the spread of the ensemble forecast (light grey represents area between 10th and 90th percentile and the darker grey area represents the spread between the 25th and 75th percentile), blue lines are the ensemble mean posterior distribution, green x are data points assimilated into the model and red * are future observations.

**Supplementary Video S2**.

**The evolution of dengue fever in Guangdong Province in 2014**. The brown dots indicate dengue cases exist at the current time. This process assumes that the duration of dengue fever cases is 7 days. Besides, the current date and the cumulative number of dengue cases are record.

